# Pan-cancer detection and typing by mining patterns in large genome-wide cell-free DNA sequencing datasets

**DOI:** 10.1101/2022.02.16.22268780

**Authors:** Huiwen Che, Tatjana Jatsenko, Liesbeth Lenaerts, Luc Dehaspe, Leen Vancoillie, Nathalie Brison, Ilse Parijs, Kris Van Den Bogaert, Daniela Fischerova, Ruben Heremans, Chiara Landolfo, Antonia Carla Testa, Adriaan Vanderstichele, Lore Liekens, Valentina Pomella, Agnieszka Wozniak, Christophe Dooms, Els Wauters, Sigrid Hatse, Kevin Punie, Patrick Neven, Hans Wildiers, Sabine Tejpar, Diether Lambrechts, An Coosemans, Dirk Timmerman, Peter Vandenberghe, Frédéric Amant, Joris Robert Vermeesch

## Abstract

**Background:** Cell-free DNA (cfDNA) analysis holds great promise for non-invasive cancer screening, diagnosis and monitoring. We hypothesized that mining the patterns of big datasets of shallow whole genome sequencing cfDNA from cancer patients could improve cancer detection.

**Methods:** By applying unsupervised clustering and supervised machine learning on large shallow whole-genome sequencing cfDNA datasets from healthy individuals (n=367), patients with different hematological (n=238) and solid malignancies (n=320), we identify cfDNA signatures that enable cancer detection and typing.

**Results:** Unsupervised clustering revealed cancer-type-specific sub-grouping. Classification using supervised machine learning model yielded an overall accuracy of 81.62% in discriminating malignant from control samples. The accuracy of disease type prediction was 85% and 70% for the hematological and solid cancers, respectively. We demonstrate the clinical utility of our approach by classifying benign from invasive and borderline adnexal masses with an AUC of 0.8656 and 0.7388, respectively.

**Conclusions:** This approach provides a generic and cost-effective strategy for non-invasive pan-cancer detection.

## Introduction

Cell-free DNA (cfDNA) is a promising non-invasive biomarker in liquid biopsy for cancer management. Shallow whole-genome sequencing (sWGS) of cfDNA can identify cancer-specific copy number aberrations (CNAs) in cancer patients (1,2). Using genome-wide cfDNA sequencing data to profile genomic imbalances, we reported that CNAs in the asymptomatic population can be indicative of incipient tumors and has potential as a cancer screening tool (3).

In addition to CNAs, sequencing of cfDNA provides a unique view on the genome-wide cfDNA fragmentation profile (4,5). CfDNA fragments carry tissue-associated nucleosome and preferred end position information (6,7), reflecting tissue-specific degradation, chromatin accessibility and nucleosome organization of its cellular origin (8,9). In healthy individuals, plasma cfDNA comprises DNA fragments that are mainly resulting from apoptotic release of DNA from the cells of hematopoietic origin (10). In plasma of cancer patients, circulating tumor DNA (ctDNA) has decreased fragment sizes and signatures of the tissue of origin (8,11). Consequently, fragmentomics is emerging as an approach to reveal cfDNA properties, broadening the potential of cfDNA as a biomarker (4,12).

Increasing availability of cfDNA sWGS data from large-scale liquid biopsy projects offer unique opportunities to explore the cfDNA profiles by machine learning. We hypothesized that mining variation between sWGS profiles may uncover distinct patterns that can be associated with different pathological or physiological states. Hence, we applied an unsupervised clustering analysis and supervised machine learning workflow, which we term GIP*Xplore*, on a large number of genome-wide sWGS cfDNA profiles from cancer patients with different hematological and solid tumors and unveiled cancer-type-specific and also shared tumor-associated signatures that are absent in healthy individuals. This approach enables accurate detection of different cancers and allows prediction of the cancer types.

## Materials and Methods

### Patients and clinical data

The study was approved by the ethical committee of the University Hospitals Leuven (S57999, S62285, S62795, S50623, S56534, S63240, S51375, S59207, S64205 and S64035). Samples and consents were obtained from healthy controls and cancer patients. Blood was collected either into Streck Cell-Free DNA BCT or Roche Cell-Free DNA Collection Tubes. Plasma was isolated via a standard, two-step centrifugation procedure and stored at -80°C. Previously published sequencing data from 260 healthy subjects (3) and 177 patients with Hodgkin’s lymphoma (13) were included in the study.

### sWGS Analysis

cfDNA was extracted from plasma using standard processing procedures and sWGS sequencing (14) (details described in the Supplemental Materials). Each sample ended with 57509 autosome bin features from standard processing. Principal component analysis (PCA) was used for dimension reduction to transform data from high dimension to low dimension. We performed the supervised learning on both the original data space and PCA transformed space and found marginal gains of performance in the majority of analyses with the original data space. As the computational time was much higher using the original data space, we used PCA features in the main analyses such that features being used in both unsupervised and supervised learning were consistent.

### GIP*Xplore*

As illustrated in **Fig. 1**, we developed GIP*Xplore* to mine sWGS cfDNA data for identification of signatures. We utilized unsupervised clustering and supervised machine learning. For unsupervised clustering, we evaluated the variance being explained from principal components (PCs) in the tumor data. Overall, the top 30, 50 and 100 PCs explained above 80%, 85% and 90% of the variance in the data, respectively. While there is no absolute optimal number of PCs to be used for further analysis, non-trivial components - 50 PCs (**Supplemental Materials**) were determined as a default number for downstream analyses in the results. The Euclidean metric was used to measure dissimilarity among samples for clustering analysis. Proximity matrix based on dissimilarity of samples was generated. The t-distributed stochastic neighbor embedding (tSNE) (15) was used to map high-dimensional data to two (or three) dimensions and to visualize the clusters. Due to the random process of tSNE, we applied Walktrap community (16) detection on the original proximity matrix for cluster assignments regardless of the presentation of tSNE visualization. In running tSNE, we set parameters perplexity of 15/30 and iteration of 10000 with exact tSNE for accuracy, and the process was repeated for 10 times with different seeds. For Walktrap, we used the parameters of 8 initial numbers of neighbors search and a walk step of 2. Clusters defined from the community detection were used for annotation. In supervised learning, PCA transformed genome-wide features were used in the machine learning model for training. PCA was performed on training data, and test data was projected on PCA space of training data for classification tasks. We measured performance by repeating the tenfold cross-validation 10 times and leave-one-out (LOO) procedures. For cross fold (CV) validation, the ROC curve and performance was calculated by averaging over 10 repeats. For classifiers, we used a support vector machine (SVM) and hyperparameters were chosen based on the grid search with a subset of the data. A separate model was trained to localize tissue of origin and LOO was used to evaluate performance characteristics. Weighted sample size was accounted for in the model for imbalanced classes.

**Fig. 1.**
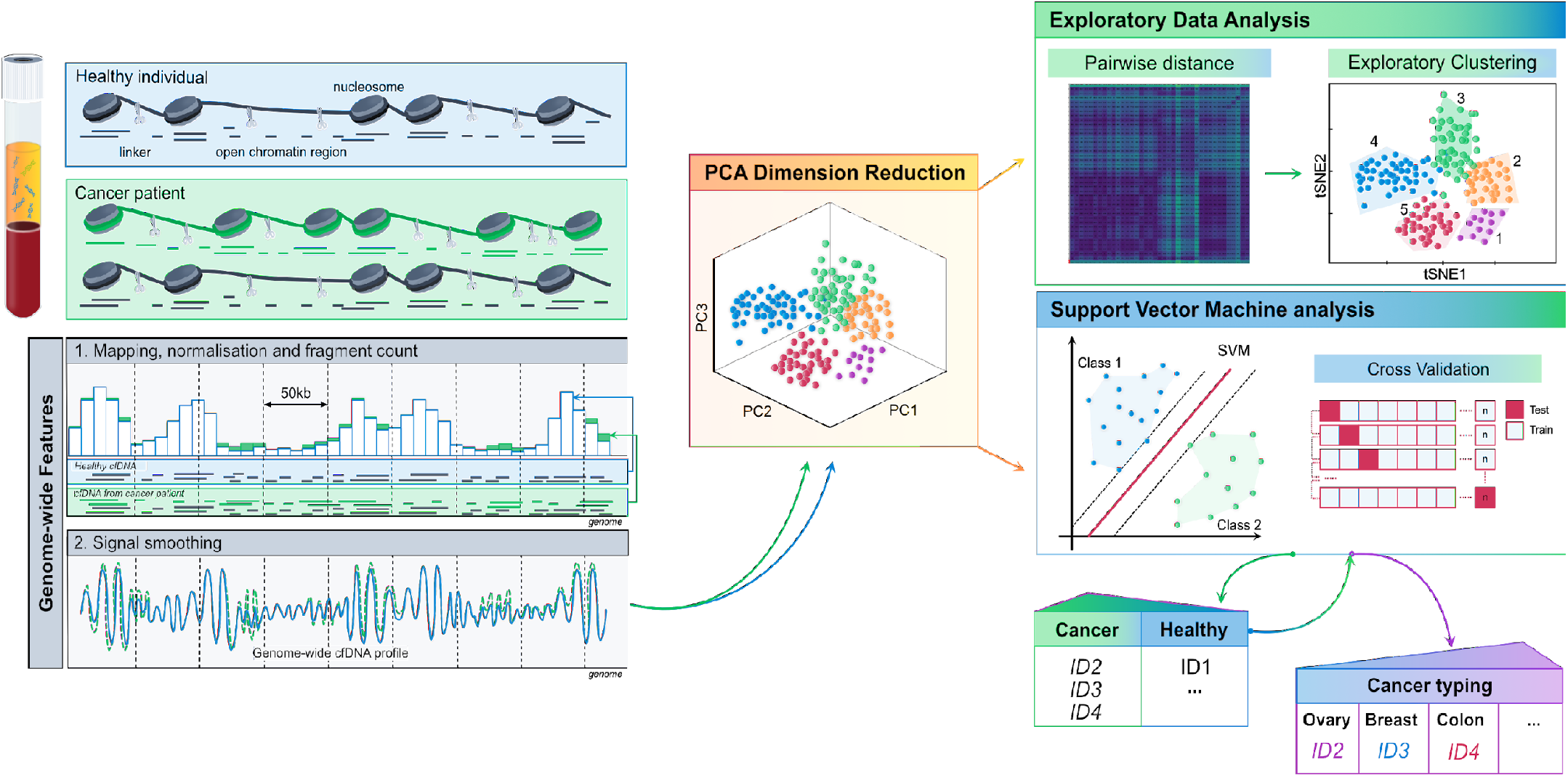
Schematic illustration of GIP*Xplore*. Plasma cfDNA in healthy individuals (blue box) comprises short nucleosome-protected DNA fragments mainly released from the cells of hematopoietic origin. In patients with cancer (green box), cfDNA is also released from the tumor. Since cfDNA fragmentation pattern is cell- or tissue-specific, sequencing and mapping of cfDNA from a patient with cancer may have differential genome-wide distribution of DNA fragments along the genome compared to a healthy one (green and blue profiles respectively). The workflow of GIP*Xplore* combines two tasks. First, explorative analysis of the high-dimensional data is performed via unsupervised clustering. Data complexity is reduced by using the first 50 linearly transformed genome-wide coverage features (non-trivial principal components, PCs) from a large number of cfDNA profiles, which are used for dataset exploration to unveil the potential biological signals or technical confounding factors based on the sub-grouping of underlying patterns that facilitate the design of the supervised models. Concurrently, classifiers are constructed to predict disease status and identify disease type to assess the use of such transformed genome-wide features as a marker for diagnostic application.

## Results

### GIP*Xplore* detects and classifies hematological malignancies with high accuracy

To assess the potential, we applied our method on a set of cfDNA samples from healthy controls (n=260) and patients with hematological malignancies that included Hodgkin’s lymphoma (HL; n=179), diffuse large B-cell lymphoma (DLBCL; n=37), multiple myeloma (MM; n=22) (**Table 1**). Walktrap community detection was performed on the dataset, and 15 clusters were defined. Visualization with the tSNE yielded separations between malignant and healthy control profiles, and the tSNE representation was largely in agreement with the clusters found by Walktrap (**Fig. 2, A**). Moreover, we observed cancer type-specific clusters. Cluster 1, 3 and 4 was exclusively composed of HL samples. Cluster 9 was enriched for DLBCL samples, and cluster 13 was specific to MM samples (**Fig. 2, B** and **Supplemental Fig. 1**).

**Table 1.**
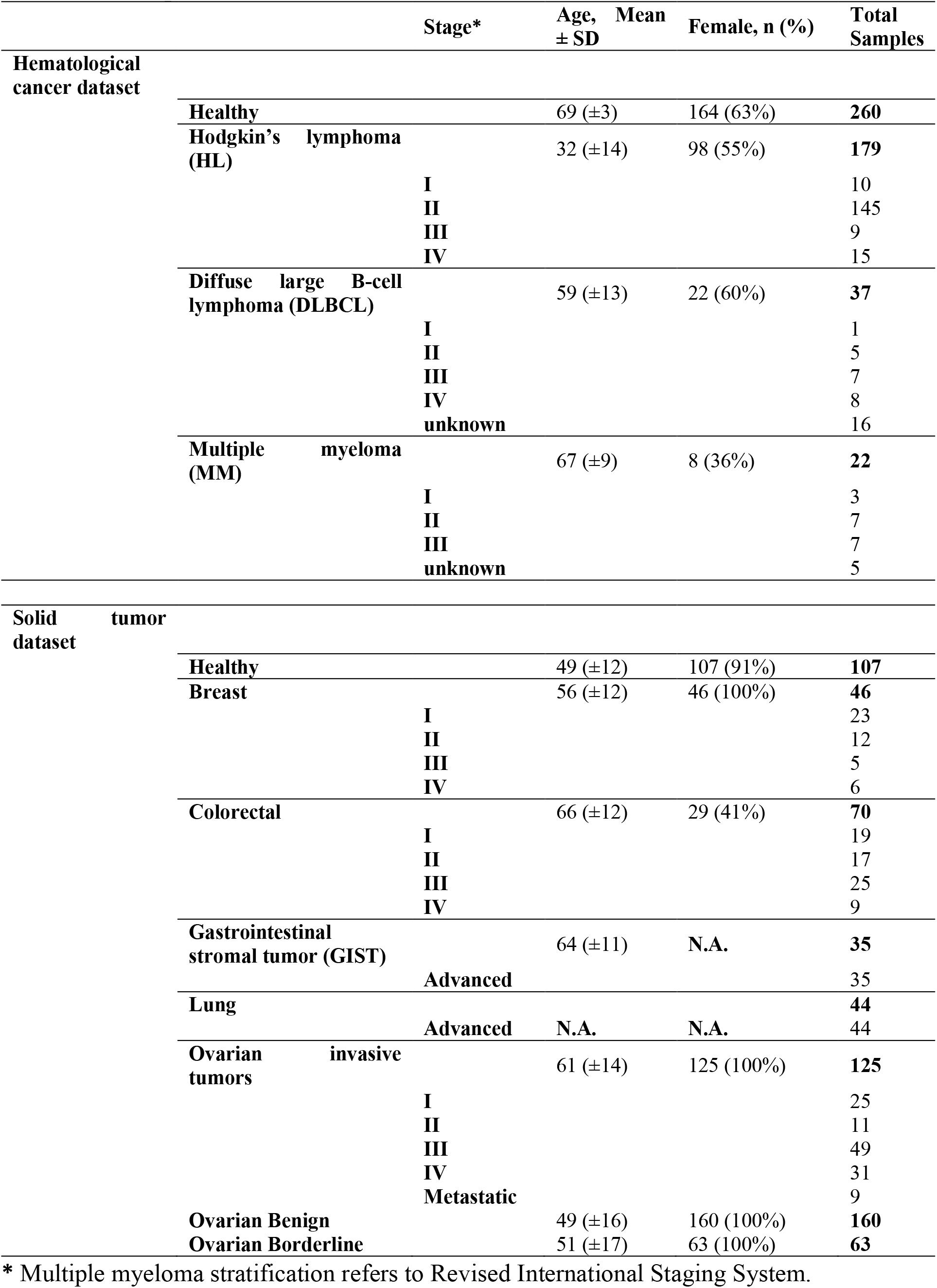
Participant and characteristics.

**Fig. 2.**
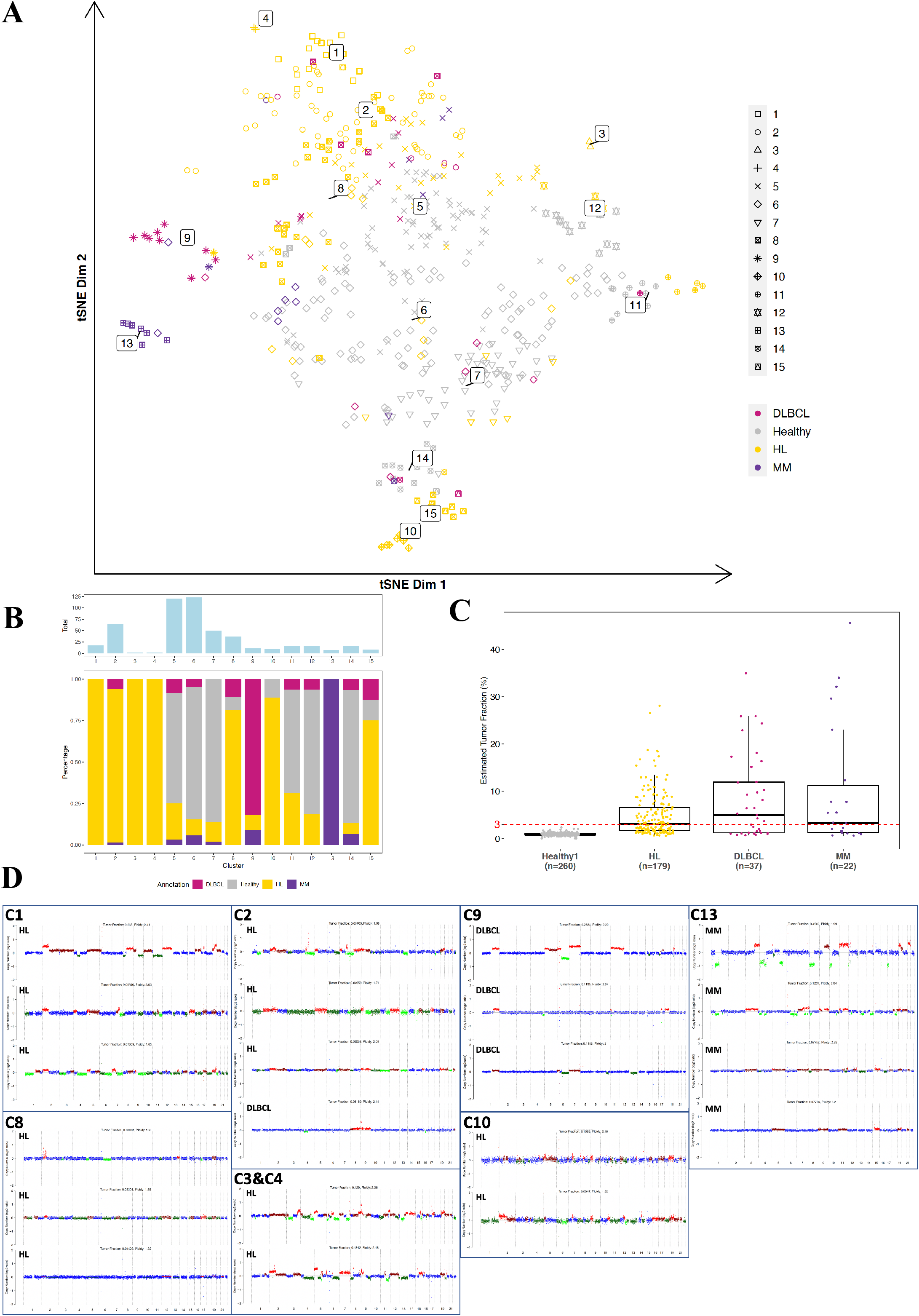
Genome-wide cfDNA profiles carry cancer type-specific patterns. **A**, Two-dimensional tSNE visualization of the clustering result. Sample type is annotated by point color and community detection resulted clusters are annotated by point shape. Cluster numbers are labeled in the center of the defined cluster. **B**, Sample distribution in each community detection defined cluster is shown. The upper bar plot shows the total number of samples grouped in each cluster and the lower bar plot depicts the proportion of each class of samples. **C**, Tumor fraction estimated using ichorCNA. Red horizontal line indicates a detection limit of 3% tumor fraction level. **D**, Examples of copy number profiles generated from ichorCNA for selected clusters. In each copy number profile, color red represents copy number gains and green represents copy number losses. The color is supposed to be interpreted together with the log ratio values to pinpoint copy number gains or losses.

In parallel, we benchmarked our method against the ichorCNA (17) algorithm for copy number profiling and tumor fraction (TF) estimation from sWGS data. IchorCNA utilizes the depth of coverage to evaluate the presence of large-scale copy number aberrations and the probabilistic model is used to infer copy number states and estimate fraction of tumor. Overall, only 52.95% of hematological cancer samples had detectable tumor-derived cfDNA levels (**Fig. 2, C, Supplemental Fig. 2** and **Supplemental Table 1**), using the 3% detection limit suggested in the ichorCNA for detecting the presence of tumor. The above-mentioned clusters 1, 3, and 4 consisted of profiles characterized by large chromosomal aberrations and high tumor load. Clusters 2 and 8 consisted of profiles from HL patients with both high and low tumor fractions, implying that the clustering was not completely CNA-driven. In particular, ten out of 65 (15.38%) lymphoma samples in cluster 2 with normal-like profiles (without detectable CNAs) grouped together with samples characterized by detectable CNAs. A less pronounced separation could be observed between clusters containing healthy controls and cluster 8, in which 76.47% (26 out of 34) malignant cases had normal-like profiles with less than 3% TFs. Nine HL samples in cluster 10 showed higher bin-to-bin log2 ratio variations and were more likely to be noisy on a genome-wide scale (**Fig. 2, D** and **Supplemental Fig. 3**). The remaining malignant cases without detectable CNAs co-localized with healthy controls. To further explore whether clustering of malignant samples would be mainly CNA-driven, we performed clustering analysis using the log2 copy ratio values produced by ichorCNA. The analysis revealed that genome-wide copy number ratios alone were less informative (**Supplemental Fig. 4**). In addition, we tested whether our method could detect underlying genome-wide changes irrespective of the presence of CNAs by restricting the clustering to the cancer samples with low TF (< 3%). The separation between some malignant and healthy samples still remained (**Supplemental Fig. 5**). Collectively, the clustering analysis on genome-wide features showed separation between malignant and healthy profiles and grouping of similar cancer type-specific profiles.

The unsupervised learning delineated cancer-associated profile changes, which suggested that a more precise prediction can be made by learning representations within different tumor types using supervised classification. Therefore, we evaluated the capability to detect cancer signals and identify cancer types with supervised learning on the hematological cohort. Both leave-one-out (LOO) and repeated 10-fold cross validation (CV) was used to assess the performance of the classifier. Incorporating transformed genome-wide features, the SVM machine learning model correctly classified 220 (out of 238) malignant cases in LOO analysis, at a sensitivity of 92.44% (95% CI: 88.31% - 95.46%) and a specificity of 98.46% (95% CI: 96.11% - 99.58%), including 170 HL, 32 DLBCL and 18 MM cfDNA samples (**Supplemental Table 1**). The remaining 18 misclassified malignant samples had normal-like profiles and clustered together with healthy controls (**Supplemental Fig. 6**). The detection sensitivity was the highest for HL (**Supplemental Table 1**). The sensitivity did not differ substantially between early (I-II) and advanced (III-IV) stages for these cancer types, though the distribution of the cases across clinical stages was unequal (**Fig. 3, A**). ROC analysis had an AUC value of 0.989 (95% CI: 0.980 – 0.998) in distinguishing malignant from healthy samples, compared to ichorCNA TF-based analysis which had an AUC of 0.929 (**Fig. 3, B)**. Repeated 10-fold CV also revealed a stable performance at an averaged AUC of 0.989 (**Supplemental Fig. 7**). As the clustering analysis demonstrated the co-localization of samples originating from the same cancer type, we then attempted to determine the accuracy of our GIP*Xplore* in cancer type classification. For this purpose, we trained the classification model using the 220 correctly predicted malignant samples. The analysis showed an overall accuracy of 85.45% (95% CI: 80.09% - 89.83%), with the highest accuracy in HL prediction (**Fig. 3, C** and **Supplemental Table 2**). Consistent with the exploratory clustering analysis, where some of the profiles from DLBCL patients colocalized together with those from HL patients, DLBCL samples were more likely to be misclassified.

**Fig. 3.**
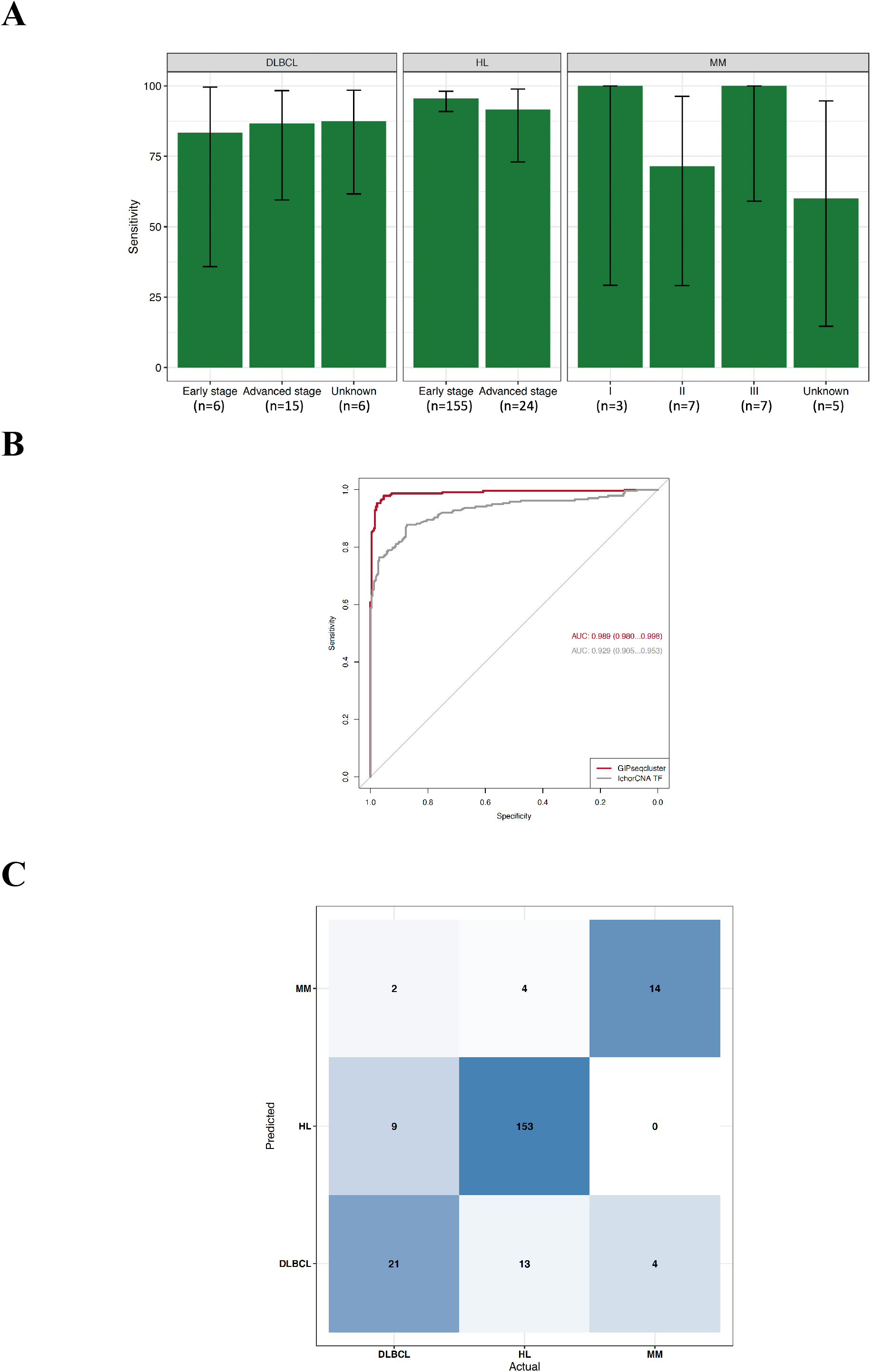
Plasma cfDNA genome-wide signatures enable hematological malignancies detection and subtype prediction. **A**, Sensitivities for detection of subtypes of hematological malignancies. Performance for early and advanced stages for DLBCL and HL are shown. Three (R-ISS) stages of MM are shown. 95% confidence interval is shown as an error bar. **B**, ROC curves for performance comparison between the genome-wide feature analysis and ichorCNA tumor fraction analysis. For the genome-wide feature analysis, decision value from SVM prediction is used to build a dynamic threshold of true and false positives. Tumor fraction values were used to construct ROC for ichorCNA analysis. **C**, Confusion matrix for tissue of origin detection in hematological tumor. The color shading represents the proportion of samples being correctly localized. The labeled numbers indicate the number of samples being classified into the class.

### GIP*Xplore* identifies and classifies different types of solid malignancies and allows disease stratification

Extending our analyses, we applied our method on a solid tumor dataset, consisting of 320 cfDNA profiles from cancer patients, and a set of 107 cfDNA profiles from healthy controls. The malignant cohort was represented by five tumor types: breast (n=46), colorectal (n=70), gastrointestinal stromal tumor (GIST; n=35), lung (n=44) and ovarian (n=125; **Table 1**). Using GIP*Xplore*, 19 clusters were identified in the solid tumor dataset (**Fig. 4, A** and **Supplemental Fig. 8**). The separations between malignant and control cfDNA profiles were less distinct compared to clustering results of the hematological cancer dataset. Clusters 4, 8, 10 and 12 were found to be cancer type-specific, in which cluster 4 was mainly enriched with ovarian cancer samples, cluster 8 was primarily consisting of cfDNA profiles from lung cancer patients, cluster 10 was GIST-specific and cluster 13 was mainly composed of colorectal samples (**Fig. 4, B**). Cluster 2, adjacent to clusters 4 and 8, was enriched with ovarian samples, although it co-localized with other tumors. Clusters 9 (mostly ovarian cancer) and 15 (intermixed cancer types) deviated from healthy and other malignant clusters. Majority of the cfDNA profiles from breast cancer patients resembled profiles from healthy controls, while one advanced stage breast cancer sample was found in cluster 8, and 2 samples from patients with advanced stage primary metastatic disease were found in cluster 2 (**Fig. 4, A** and **B**).

**Fig. 4.**
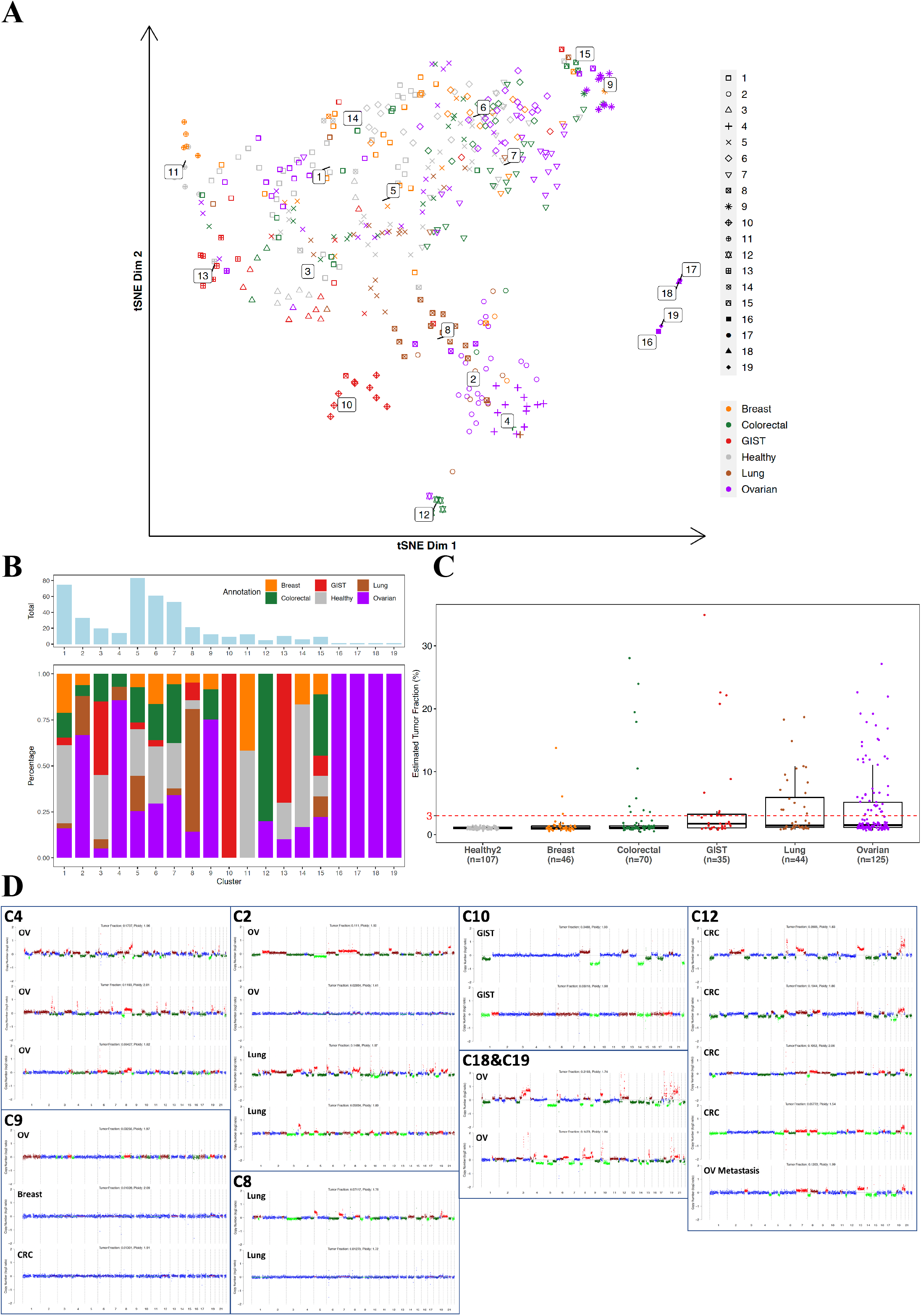
Clustering analysis elucidates profile representations in solid tumors. **A**, Two-dimensional tSNE visualization of solid tumor dataset clustering result. Sample type is annotated by point color and community detection resulted clusters are annotated by point shape. Cluster numbers are labeled in the center of the defined cluster. **B**, Sample distribution in each community detection defined cluster is shown. The upper bar plot shows the total number of samples grouped in each cluster and the lower bar plot depicts the proportion of each class of samples. **C**, Tumor fraction estimation for indicated types of solid tumors. Red horizontal line indicates a detection limit of 3% tumor fraction level. **D**, Examples of copy number profiles generated from ichorCNA for selected clusters.

Compared with the hematological cancer dataset, ctDNA levels estimated by ichorCNA were generally lower in the solid malignant cohort (**Fig. 4, C**). Tumor fraction varied among different types of cancer and increased with the stage (**Supplemental Fig. 9**). The malignant cases with detectable CNAs and therefore higher TF were more likely to separate from the healthy controls (**Supplemental Fig. 10**). Cluster 4 contained ovarian cancer samples with detectable chromosome instability. Among lung cancer profiles in cluster 8, 64.29% (9 out of 14) had detectable CNAs. Clusters 16 to 19 included four ovarian samples with high chromosomal instability that greatly deviated from other profiles. Overall, in clusters 9 and 15, profiles tended to be noisy, without clear CNAs (**Fig. 4, D**), however they deviated from healthy control and other malignant clusters (**Fig. 4, A)**. When using the log2 copy ratio profiles from the CNA analysis to investigate whether the sub-grouping of cfDNA profiles was driven by CNAs, cancer type-specific clustering patterns were diminished (**Supplemental Fig. 11**). When restricting the clustering analysis to samples with TF lower than 3%, samples from clusters 9 and 15 still showed deviations (**Fig. 4, A** clusters 8 and 9) from normal profiles (**Supplemental Fig. 12**).

We next investigated whether supervised learning using genome-wide features can enhance the detection of solid malignancy signals in sWGS cfDNA data. Classification of samples as either healthy or malignant (107 healthy controls and 320 malignancies) was performed using the SVM model, with performance estimated by LOO and repeated 10-fold CV. With an overall accuracy of 65.34%, we correctly detected 177 out of 320 cancer profiles (55.31% sensitivity, 95% CI: 49.68% - 60.84%), at a specificity of 95%. Performance in individual tumor types ranged from 15.22% (95% CI: 6.34% - 28.87%) for classifying breast cancer to 80.00% (95% CI: 63.06% - 91.56%) for GIST (**Supplemental Table 3**). Stage of the disease affected the detection, with a sensitivity of 26.17% (95% CI: 18.15% - 35.55%) in the early stage (I-II) *versus* 69.95% (95% CI: 63.31% - 76.03%) in the advanced stage (III-IV). In individual tumor types, it remained true that higher sensitivities were found for the advanced stages than for the early-stage diseases (**Fig. 5, A**). Colorectal cancer was an exception as sensitivities were almost the same for early and advanced cancer stages. Misclassified malignant samples had low tumor fraction, which potentially restricted the detection of underlying tumor-specific patterns (**Supplemental Fig. 13**). We could distinguish malignancy from healthy samples with an AUC of 0.827 (95% CI: 0.787 – 0.867), which again was superior to ichorCNA TF-based analysis (0.733 AUC, 95% CI: 0.687 – 0.780; **Fig. 5, B** and **Supplemental Fig. 14**). Subsequently, we explored the potential of our GIP*Xplore* method for tumor classification. When performing tumor type-specific prediction with the 171 correctly predicted primary tumor samples, the LOO validation resulted in a 69.01% (95% CI: 61.49%-75.84%) overall accuracy. Highest sensitivities (>70%) were obtained for cfDNA samples from ovarian cancer and GIST patients. At the same time, ovarian and colorectal tumor cfDNA profiles were more likely to be misassigned to each other (**Fig. 5, C** and **Supplemental Table 4**).

**Fig. 5.**
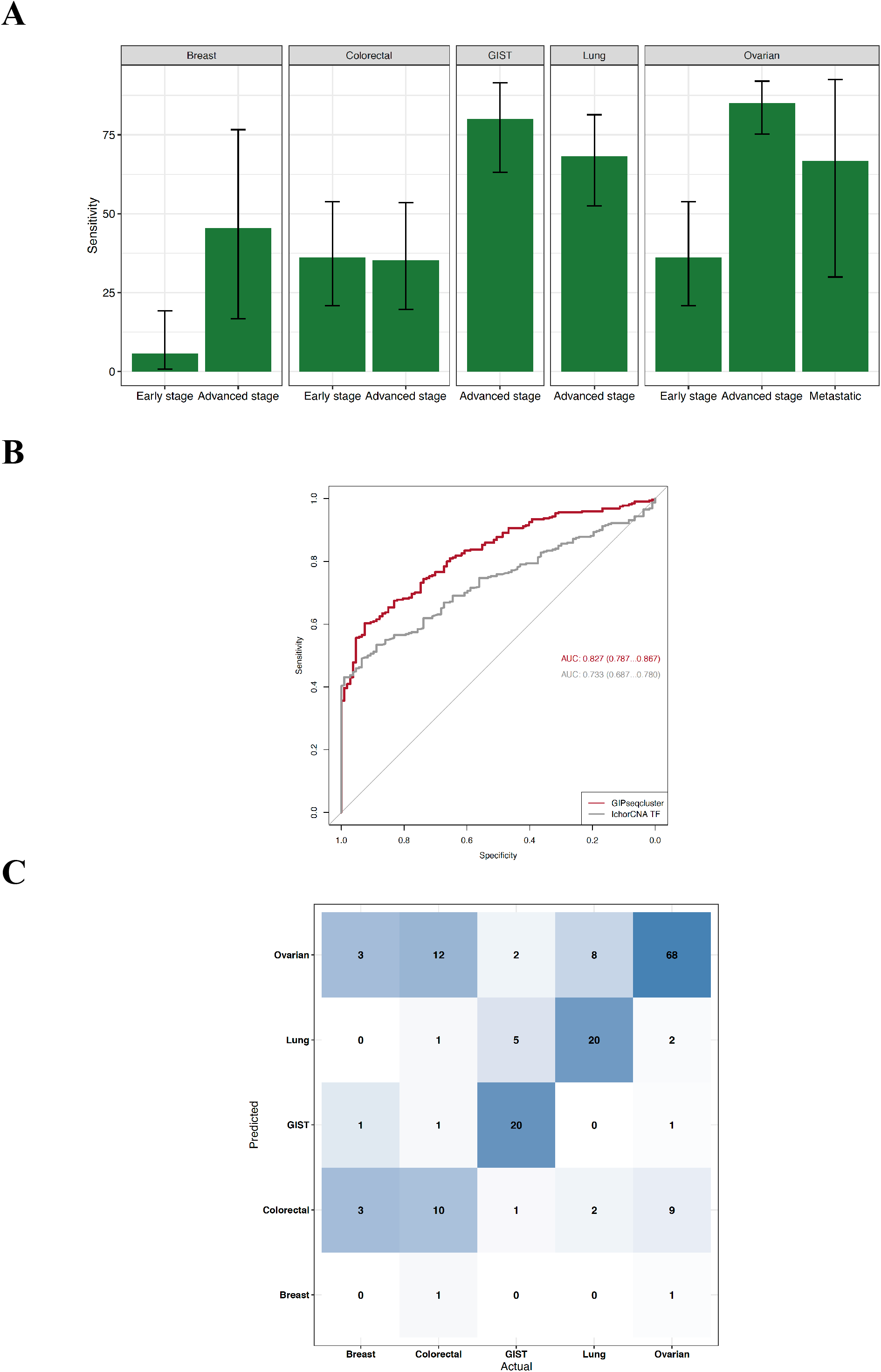
Malignancy detection and typing in solid tumors. **A**, Sensitivities for detection of different types of solid malignancies detection. Performance for detection of early and advanced stages of disease is shown. **B**, ROC curves for performance comparison between the genome-wide feature analysis and ichorCNA tumor fraction analysis. **C**, Confusion matrix for tissue of origin detection in solid malignancies. The color shading represents the proportion of samples being correctly localized. The labeled numbers indicate the number of samples being classified into the class.

Moreover, among this solid malignant cohort, we had nine cfDNA samples from patients with ovarian metastases, of which four patients had gastrointestinal primary site, one lymphoma, one leiomyosarcoma, one uterine origin, and the remaining two had Krukenberg tumors. Annotation of these nine cases on the tSNE plot showed that metastatic profiles could resemble profiles of either the primary tumor or the distant site (**Supplemental Fig. 15, A**). Applying the type-specific classifier to the six metastatic cases that were predicted as malignant cases by the malignancy classifier, the case with gastrointestinal origin that was co-clustered with colorectal samples was classified to be colorectal class. Two out of three metastatic cases that were identified in intermixed clusters of lung and ovarian tumors were predicted to be lung class and the other one was assigned to ovarian class. The additional two cases identified by the classifier were classified to the ovarian and colorectal classes, respectively (**Supplemental Fig. 15, B**).

### Accurate classification of benign from invasive and borderline adnexal masses may improve clinical management

In addition to the invasive ovarian tumor samples, our cohort contained 160 benign and 63 borderline ovarian samples. To assess the potential utility of the method for ovarian cancer management, we analyzed the ovarian tumor cohort independently by performing clustering analysis and building the ovarian-specific classifier to differentiate benign from malignant adnexal masses. Benign and borderline samples were less likely to have detectable ctDNA levels (**Supplemental Fig. 16**). In the clustering analysis, 35 invasive samples formed a distinct group in cluster 1. In clusters 6-9, common patterns were found for invasive, benign and borderline samples, although they remained distinct from controls (**Supplemental Fig. 17**). The classification analysis exhibited an AUC of 0.8656 (95% CI: 0.7761 – 0.8689) in discriminating benign from invasive samples, and an AUC of 0.7388 (95% CI: 0.6857 – 0.7920) in discriminating benign from borderline and invasive samples (**Supplemental Fig. 1** and **19**).

## DISCUSSION

We present a generic approach for cancer identification and classification by mapping genome-wide cfDNA signatures, without prior knowledge of genetic alterations or predefined signatures in the sequencing data. The unsupervised clustering allows the discovery of hidden genome-wide patterns, and the supervised learning model can be trained to detect such underlying signatures. This method can be used to classify cfDNA samples by matching to existing datasets and has the potential to be used as a pan-cancer assay for detection and typing of multiple cancers from one blood draw.

Current sWGS cfDNA analyses mainly focus on the detection of somatic CNAs (17–19). These methods are blind to events that involve copy neutral abnormalities. Our approach also differs from the previous method that classified tumor types based on selected CNAs, and in which normal-like profiles were incapable of tumor classification (20). We demonstrate that even profiles without detectable CNAs carry informative and discriminative patterns in sWGS data. Different recent studies have utilized methylation, transcription factor binding, fragment lengths, or cfChIP-seq for cancer detection (4,12,21–25). While these studies have important implications and show cfDNA as a promising biomarker, they require more specific workup and/or deeper sequencing. In contrast, analysis of sWGS data can be easily adapted in clinical settings and complement CNA analysis. By mapping differences among the cfDNA profiles, shared abnormality patterns are captured.

To date, Liu *et al*. have reported the largest population-level cfDNA methylation study for multi-cancer detection, in which the targeted methylation analysis of cfDNA enabled detection of more than 50 cancer types at a sensitivity of 54.9% and at a specificity of 99% (21). This test was refined and validated in an independent follow-up study, with an overall sensitivity of 51.5% at 99.5% specificity was reported (26). In line with these findings, we estimate the combined sensitivity of 68.03% at above 95% specificity for the hematological and solid cancer cohorts. Performance for cancer signal detection varied among the different cancer types and stages. The prediction accuracy was highest for hematological malignancies and lowest for breast cancer. Shedding of the ctDNA from breast cancer is known to be low (27,28). Also, our cohort had an over-representation of early-stage cancers, with 50% of the samples from stage I. Apart from potential screening applications, we also demonstrated that GIP*Xplore* could be used for risk stratification and management of a specific cancer type. Discrimination between malignant, borderline and benign masses at diagnosis is of critical importance to improve patient management (29,30).

The accuracy of tumor type-specific prediction might depend on the intrinsic tumor characteristics. For example, DLBCL being more heterogeneous on molecular level (31,32), had lower classification accuracy than HL and MM. The subtype of colorectal and ovarian tumors is of similar cellular origin, and histological subtypes can be hard to distinguish (33– 35), which might be a reason for misclassification amongst the two cancer types. The identification of the origin of some metastases, suggests the method may allow the identification of unknown primary cancers. The metastatic cases were classified into profiles of its primary or distant sites, possibly reflecting changes during the metastatic progression or dynamic tumor DNA shedding from tumor tissues (36–38).

Interestingly, besides tumor type- or aberration-specific subgroups, our analysis revealed the presence of additional clusters that segregated from healthy controls (**Fig. 4, B** and **Supplemental Fig. 17**). Though the origin of such segregations remains unknown, we hypothesize the method provides a system-wide insight, potentially reflecting (patho)physiological conditions of these individuals. Dynamic cellular responses and malignant cell proliferation with active involvement of immune response during (early) carcinogenesis might lead to the observed common changes in cfDNA composition across different cancer types (39,40). Therefore, it is possible that our analysis detected tumor-driven immune or other biological responses or states.

GIP*Xplore* provides an unbiased genome-wide scan of cfDNA profiles. However, it also has some limitations. Increasing the sequencing depth might improve detection of disease-specific cfDNA patterns and improve the sensitivity of our methodology further. The data presented here has a larger proportion of HL and ovarian cancer samples and is limited in the number of different cancer types, which may affect the aggregated sensitivity and distort tumor typing accuracies. We foresee that expanding the breadth of the evaluated cancer types may improve prediction of tissue/cell origin and facilitate a deeper understanding of cfDNA in the context of tumors. Increasing the range of physiological states and diseases that are relevant for these tumor samples will be essential to fully interrogate the potential and limitations of our approach. The approach may also be further broadened to project and embed new treatment or follow-up data for cancer prognosis and monitoring.

In summary, we have extended the scope of cfDNA analysis, allowing cost-effective identification of genome-wide cancer-(type-)specific signatures from shallow sequencing data, allowing improved discrimination between profiles from cancer patients and healthy individuals. This study lays the foundation for enhanced genomic characterization of cfDNA that can be used for improved cancer management. We foresee that the method can be scaled up for detection of multiple pathological conditions.

## Data Availability

All data produced in the present study are available upon reasonable request to the authors.
Processed alignments of sequencing data are archived to ArrayExpress (https://www.ebi.ac.uk/arrayexpress/) with unrestricted access under accession number E-MTAB-10934. Code will be available upon request.

## Acknowledgements

We would like to acknowledge the patients and blood donors. We would like to thank Gitte Thirion and Annick Van den Broeck for the collection of samples and the extraction of ctDNA, Kate Stanley for helpful suggestions for the manuscript.

## Author contributions

HC, TJ, LL, LD, FA and JRV conceptualized and designed the study. KP, AW, EW, AC, PN, ST, DT, PV, HW and FA provided clinical samples and patient data. LL, LV, NB, IP, KVDB, CD, DF, RH, SH, CL, L.Liekens, VP, ACT, AV and AW carried out clinical sample procurement and processing. LV, NB, IP and KVDB coordinated sequencing of cell-free DNA. TJ and LL conducted project coordination and administration. HC and LD performed bioinformatics analysis of sWGS data. HC, TJ, LL, LD, KP, AW, EW, AC, DL, ST, DT, PV, FA and JRV contributed to the interpretation of results. HC, TJ and JRV wrote the manuscript; all co-authors reviewed the manuscript.

## Data and materials availability

Processed alignments of sequencing data are archived to ArrayExpress (https://www.ebi.ac.uk/arrayexpress/) with unrestricted access under accession number E-MTAB-10934. Code will be available upon request. All other materials associated with this study are present in the paper or the Supplementary Materials.

