## supplemental figures for "Pan-cancer detection and typing by mining patterns in large genome-wide cell-free DNA sequencing datasets"

### Supplemental Materials

#### Cell free DNA extraction and shallow whole-genome sequencing

cfDNA was extracted from 2 (to 4) ml plasma, using the QIAamp circulating nucleic acid Kit (Qiagen, Hilden, Germany; manual extraction), Mag-Bind cfDNA Kit (Omega Bio-tek), or the Maxwell HT ccfDNA kit (Promega, Madison; automated procedure). DNA sequencing libraries were prepared using the Truseq Chip (Illumina) or KAPA HyperPrep (Roche Diagnostics) kits. Whole-genome sequencing was carried out on a HiSeq2000 (17.30% of the samples), HiSeq2500 (23.52%), HiSeq4000 (52.87%) or NovaSeq (6.31%) (Illumina) generating single-end 36 or 50 bp or pair-end 2x101 bp reads. After filtering, 2 – 25 million reads (0.02-0.25-fold coverage), with a median number of 9.16 million reads, were obtained for each sample.

#### Sequencing data processing

Sequencing data from all samples went through the same processing procedures. Raw reads were clipped to single end 36 bp and were mapped to the human reference genome GRCh38 using BWA (v0.7.11). Duplicated reads were removed by Picard MarkDuplicates (the Broad Institute). Low quality mapping reads (< 20) and secondary alignment were removed by Samtools (v1.9). Reads mapped to blacklist regions (centromere and N-regions defined from UCSC genome and the GRC reference) were excluded from the downstream analysis. Autosomes were partitioned into non-overlapping 50-kb bins. Read counts per bin were calculated and normalized by median bin count. Locally weighted scatterplot smoothing (LOESS) regression analysis was applied to correct the biases in counts attributed to the GC content. To denoise and smooth the bin count, a sliding window of 20 with a stride per bin was applied and a mean normalized count per window was calculated.

#### Genome-wide features

During the exploratory data analysis phase, we tested different bin sizes for genome-wide feature estimation. We generated profiles with 10kb, 50kb and 100kb bins. 10kb bins can result in more than 300,000 feature vectors, which requires more computations in downstream analysis. 100kb bins profiles were less performed than 50kb bins-based profiles in discriminating between pregnant and non-pregnant subjects (data not shown). Therefore, 50kb bin was used as the unit feature and selected for further analyses.

#### Determining the number of non-trivial principal components

The scree plot showed (**Supplemental Fig. 20**) the cumulative variance being explained in PCA using hematological and solid tumor dataset, respectively. For datasets involving cfDNA samples from cancer patients (hematological and solid tumor cohorts), the cumulative variance curve hit the ‘elbow’ at the range of 20-30 PCs, and after 50 PCs, the cumulative variance being explained in the dataset increased in a diminished manner. To further determine the number of non-trivial components to keep, we compared the observed *n*th eigenvalue ($\lambda_1 \geq \lambda_2 \geq ... \geq \lambda_n$) with the distribution for the *n*th eigenvalue found in data using randomized permutation test. The eigenvalues were obtained from the input data. 5000 replicates of the input data were then generated, and in each replicate, each count feature was randomly shuffled. PCA was applied on the shuffled replicates and corresponding eigenvalues were obtained. P-values for the components were computed under the null hypothesis – the *n*th component is trivial. The procedure was repeated for 5 times with stratified random sampling to 90% of the data. For the two datasets, the cutoff for non-trivial components were around 39 and 35 (**Supplemental Fig. 21**). Taken the analyses together and accounting for subtle signals, we proceeded with 50 components for further analyses.

**Distance metrics**
Non-trivial principal components (n=50) were used for profile similarity measure and tSNE analysis. We computed pairwise distances among samples in the datasets, using the Euclidean, Manhattan, Pearson's and Spearman's distances (**Supplemental Fig. 22-23**). Euclidean and Manhattan distances preserve the scale differences, while correlation-based distances only consider the association. Euclidean and Manhattan distances showed similar patterns. We selected Euclidean distance as the metric for dissimilarity measurement in the work.

**Walktrap community detection**
Results from different iterations of tSNE showed a high stability. While tSNE reflects reasonable coherent patterns in high dimensional space, it could be possible to over-interpret clusters that appear coherent in the tSNE embeddings. To have a more deterministic outcome on defining the clusters, the Walktrap method was used to perform random walks on the constructed network graph to identify clusters/communities, as the walks are more likely to stay in the densely connected clusters[1]. The process begins on a selected node (entity) and moves to another node chosen randomly and uniformly from its neighbors, and then proceeds to a next node in the same way, with the number of steps specified as the walk[2]. The network was constructed from the Euclidean distance matrix, and the nearest number of 8 was used to build edges, weighting on the distance. Subsequently, the Walktrap was used to partition the built network. The parameter - length of walk can result in different levels of granularity, with the length to be short enough to be meaningful but long enough to gather community information. A walk step length of 2 was specified here to start the graph with high resolution. While there is no definitive answer to the optimal number of clusters, we used the modularity metric to estimate the potential optimal partition of the communities. Modularity measures the strength of the community division and quantifies the density of links inside the communities compared to links between the communities without the use of ground truth[3]. As the Walktrap produces hierarchical divisions, we calculated the overall modularity of the community structures at different cut points[4]. The maximum cut point corresponds to the number of nodes. The cut at which the maximum modularity value was returned was selected as a final output for cluster numbers/community partitioning (**Supplemental Fig. 24-25**).

**Classifiers for cancer profile prediction**
To train a classifier to distinguish healthy from cancer profiles, the e1071 and caret R packages were used. Radial kernel was used in support vector machines. Hyperparameters – gamma and cost, was tuned by the grid search on a randomly selected 90% of the data. The selected hyperparameters were used in tenfold cross validation and leave-one-out validation. To estimate prediction performance, we used both tenfold cross-validation and leave-one-out validation. To take the randomization into account, we repeated the procedure ten times in tenfold cross-validation. The top 50 principal components, performed only on the training data in each cross-validation run, were retained to build the prediction model. Different weights that were inversely proportional to the class distribution were assigned to classes in model training to address the imbalanced class sizes. Test data in each cross-validation run was projected onto the PCA space of the training set and the top 50 PCs after transformation were fed into the trained model for prediction. For prediction, decision value was obtained and later was used in the ROC analysis.

Additionally, for cancer profile prediction, we assessed performance using the original genome-wide features without PCA transformation. With the same model building procedures, the estimated performances for identifying cancer signals using untransformed genome-wide features (**Supplemental Fig. 26**) were comparable to those of using principal components, but with more computational time required.

#### Classifier for tumor type prediction

We trained a separate model for samples that were correctly predicted as cancer profiles to localize the tissue of origin. Considering small sample sizes for some tumor classes, performance characteristics were assessed using leave-one-out validation. The multi-class SVM model training procedures were the same as those in the cancer prediction. The multi-class model trained using all correctly predicted cancer profiles was applied on the metastatic cases to predict tumor type. Each metastatic sample was projected on the PCA space of the training data and the class was assigned.

#### Investigation of technical variations

Inspection of the data using clustering has revealed that pre-analytical factors, *i.e.* usage of different library preparation kits, could result in cluster separations (**Supplemental Fig. 27, B**). Hence, to avoid the bias, we analyzed the data of hematological and solid tumor cohorts separately. Other potential pre-analytical factors, including sequencing batch, age and sex, and they did not indicate an apparent confounding effect on the clustering of cfDNA samples (**Supplemental Fig. 27, C-E**). As such, unsupervised clustering can serve also as an initial quality control step to identify bias or confounding factors from unseen samples. The ability to detect technical bias can be leveraged to improve downstream analysis by minimizing technical variations.

Two kits were used for library preparation across all datasets in this study. The hematological cohort was prepared using the TruSeq ChIP kit (Illumina), while the rest of the samples were processed with the KAPA HyperPrep DNA Library Preparation kit (Roche Diagnostics). Samples that underwent different library preparation methods were apparently identified as separated clusters (**Supplementary Fig. 27, B**). The profile changes were systematic shifts, potentially due to GC content distribution (**Supplementary Fig. 28**). It is known that depending on the polymerase enzyme used in the library preparation kit, fragments with low or high GC content are likely to be amplified with different efficiency, resulting in biased read accuracy and/or distribution of the sequencing reads across the genome[5], which in the end could affect the analysis. To further check that the forming of the separate technical clusters was mainly due to the library preparation kits, we annotated the clusters with sequencing batches and found no batch-specific clusters (**Supplementary Fig. 27, C**). The sequencing batch did not seem to have an apparent confounding effect on the clustering. Additionally, plasma cfDNA from 11 healthy and 5 ovarian individuals were re-sequenced in different batches. Though some variations from batches could be noted, the profiles for two samples from the same individuals were comparably correlated (median spearman correlation of 0.9587), with greater technical reproducibility than randomly selected sample pairs (median spearman correlation of 0.9377). We could effectively monitor systematic technical changes of the profiles by the clustering. Ruling out such technical differences is essential for unbiased investigation of biological signals, and it could benefit more accurate detection by cancelling out the background noise.

#### IchorCNA analysis

For copy number analysis, additional 100 healthy controls (50 females and 50 males) that were generated in line with other samples were used as a reference panel. Deduplicated reads with quality higher than 20 were extracted with a window size of 500kb. Following parameters were used to call copy number profiles.

--ploidy \"c(2)\" --normal \"c(0.75,0.85,0.9,0.95,0.97,0.99) --maxCN 5 --includeHOMD True --chrs \"c(1:22)\" --chrTrain \"c(1:18,20:22)\" --estimateNormal True --estimatePloidy True --estimateScPrevalence False --scStates \"c()\" --txnE 0.9999 --txnStrength 10000

Optimal solution estimated by expectation-maximization convergence was used to obtain tumor fraction. For plotting, we customized the color scheme for the bins (shown as dots) by using only the blue color when the tumor fraction is below 3%. For profiles with tumor fraction greater than 3%, the original color scheme (green for deletions and red for amplifications) was kept. For segments (shown as horizontal segments across dots) coloring, the original color code was applied, regardless of tumor fraction levels. Though we observed technical difference when performing clustering on genome-wide profiles, such technical bias had marginal effects on the ichorCNA tumor fraction estimation (**Supplemental Fig. 29**). Hence, one common reference panel was used in ichorCNA analysis.

**Statistical analysis**

Statistical analyses were performed using R version 3.6.0. For principal component analysis, eigenvalue decomposition was performed with the R prcomp function. The R packages caret and e1071 were used to implement the classifiers for cancer prediction and typing. Confidence intervals for accuracy from the model output were obtained with the pROC package. Confidence intervals for sensitives and specificities were calculated using the exact Clopper-Pearson confidence intervals.

**Supplemental Figure 1. Clustering of the cfDNA profiles from hematological malignancy dataset.**

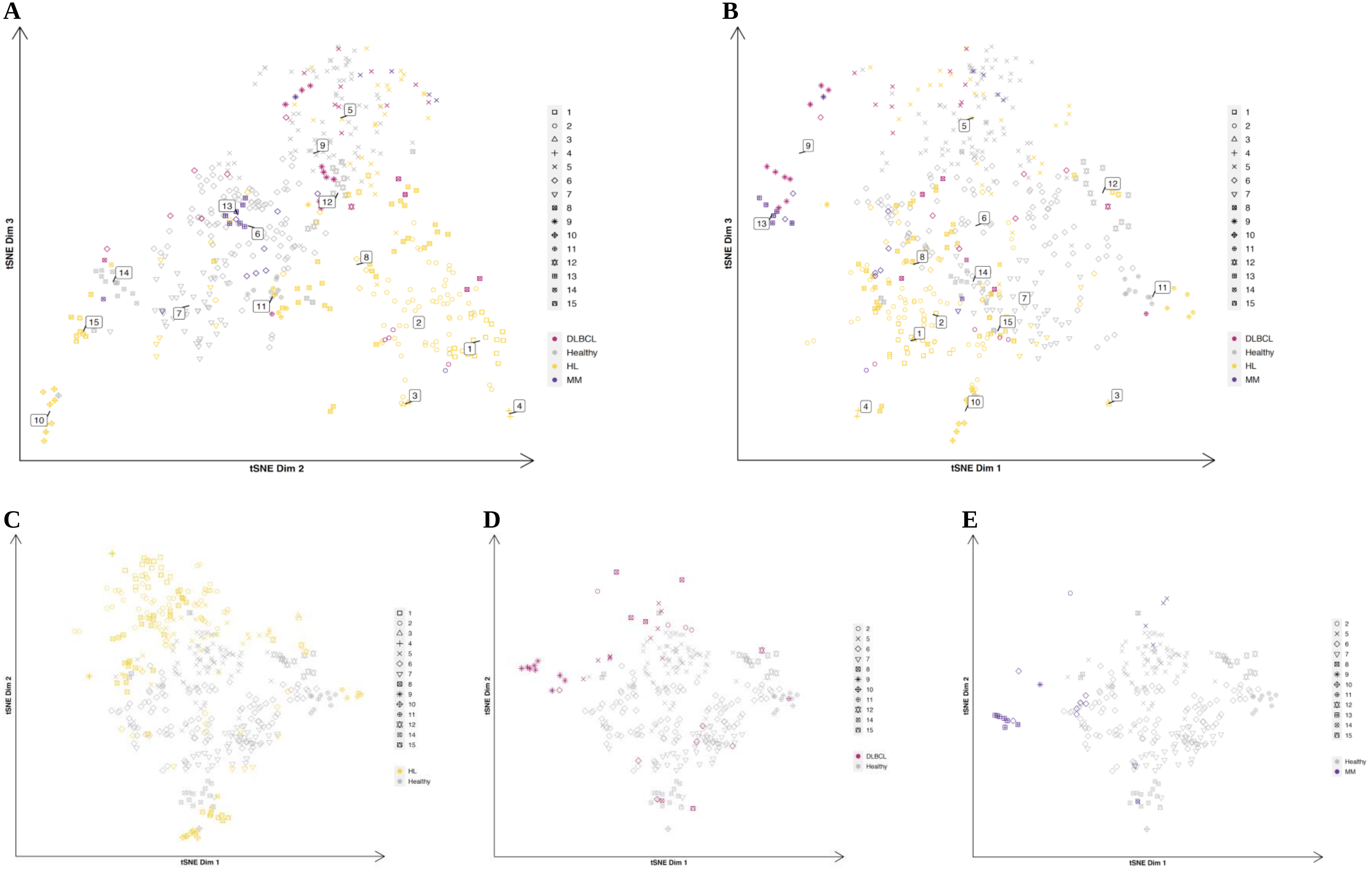

**Supp. Fig. 1.** 2D tSNE plots for the cfDNA profiles from hematological dataset. **A,** Visualization of dimensions 2 and 3. **B,** Visualization of dimensions 1 and 3. **C-D**, Cancer type-specific view of Hodgkin's lymphoma (HL, yellow), diffuse large B-cell lymphoma (DLBCL, pink), Multiple myeloma (MM, purple), respectively.

**Supplemental Figure 2. ichorCNA-estimated tumor fraction for hematological malignancies.**

**A**

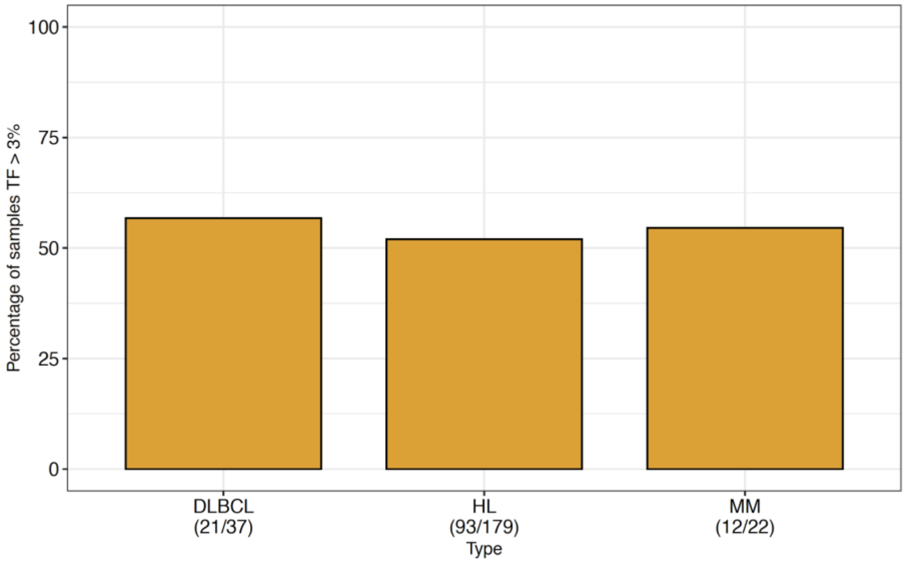


**B**

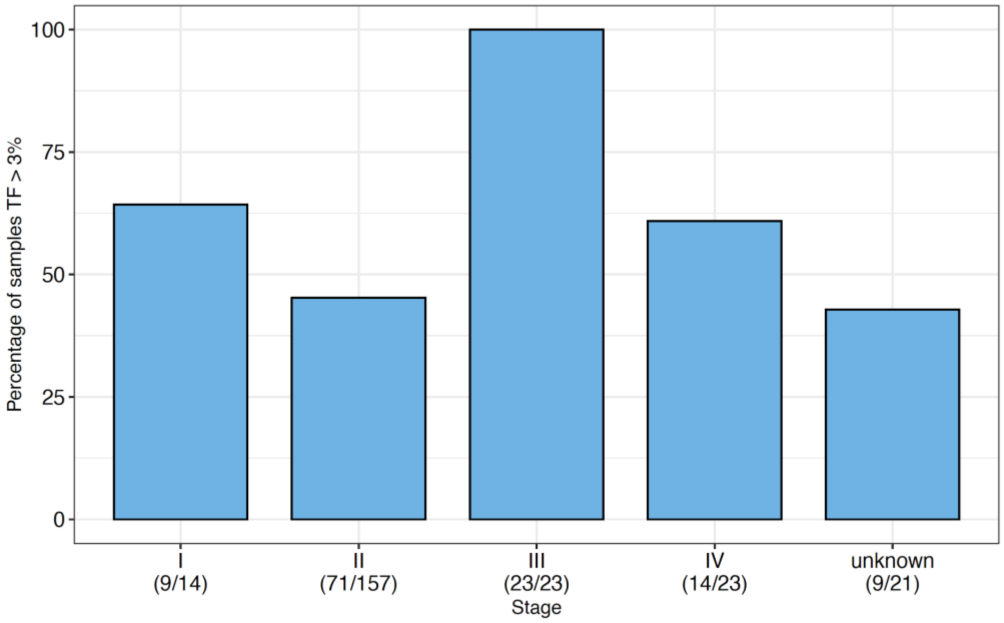


**Supp. Fig. 2. A,** Proportions of samples with tumor fraction (TF) above 3% for each type of hematological malignancies. **B,** Proportions of samples with tumor fraction above 3% across different stages.

**Supplemental Figure 3. Tumor fraction distribution across defined hematological clusters.**

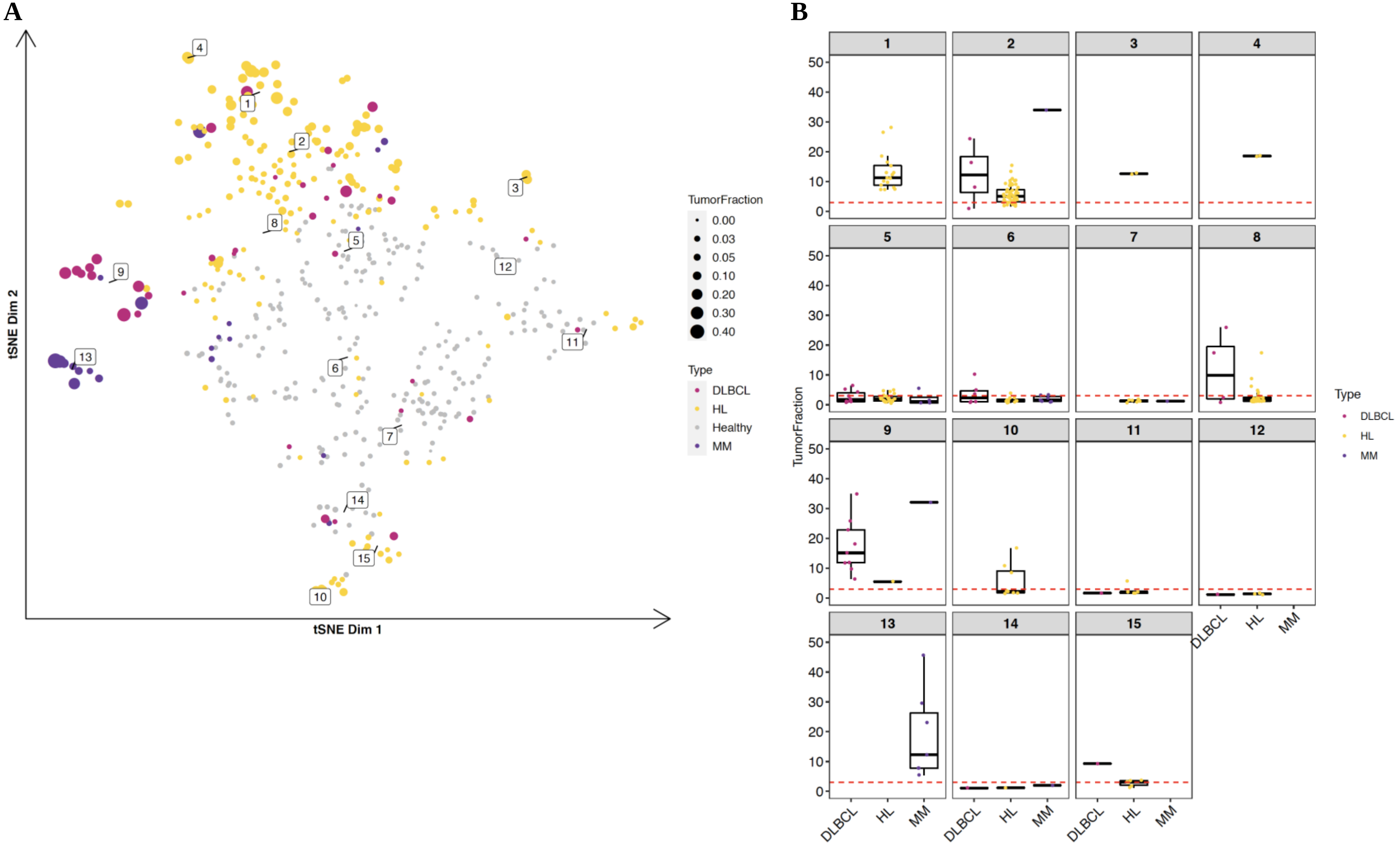


**Supp. Fig. 3.** Tumor fraction distribution across clusters of the hematological dataset. **A,** Samples with annotated tumor fraction and malignancy type annotation on tSNE visualization. The size of the point indicates the tumor fraction level - the larger the point, the higher the tumor fraction. Color indicates cancer type. **B,** Cluster-specific tumor fraction distribution for each hematological cancer type.

**Supplemental Figure 4. Clustering on the log2 copy ratio profiles of hematological dataset.**

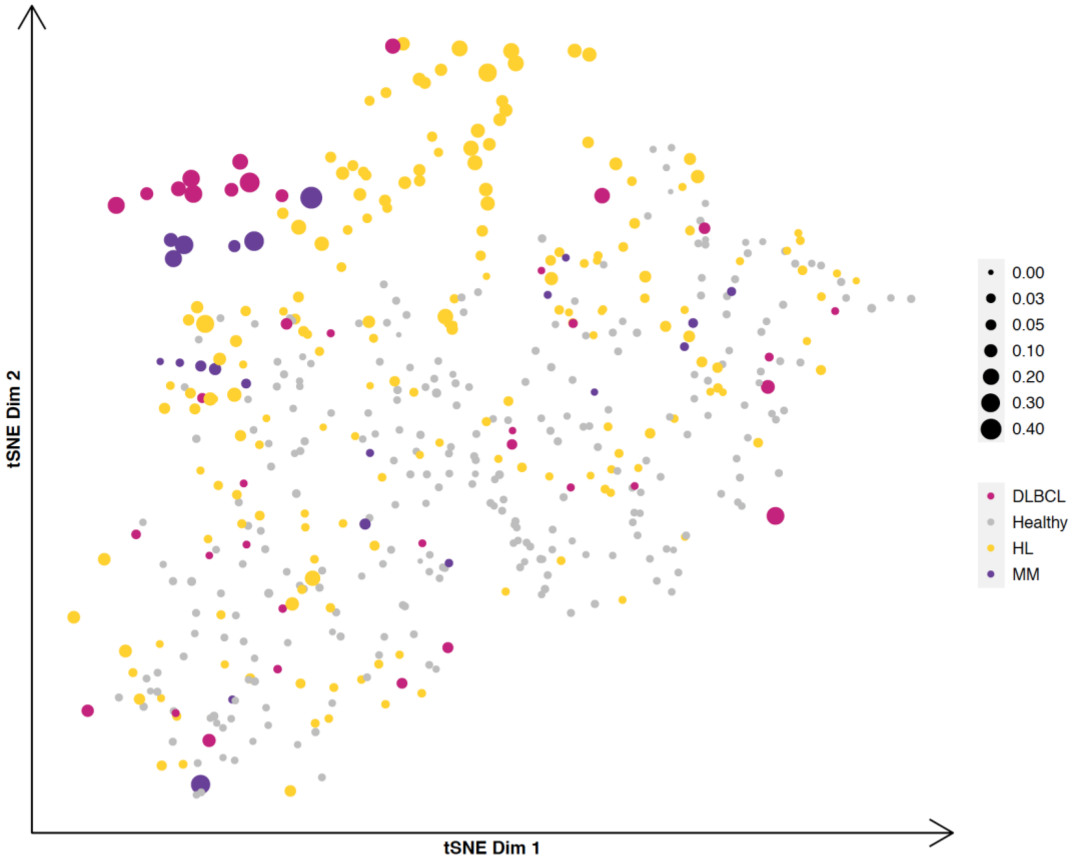


**Supp. Fig. 4.** 2D visualization of clustering using the genome-wide log2 copy ratio values from the ichorCNA analysis**.** Spearman's correlation distance was used for embedding. Euclidean distance is likely to exaggerate the discrepancy of outliers in few dimensions. In the respect to log ratio values, the scales can be of high variations, and thus the Euclidean distance loses robustness

**Supplemental Figure 5. Clustering on cfDNA samples with low TF (<3%) from hematological cancer cohort.**

**a**

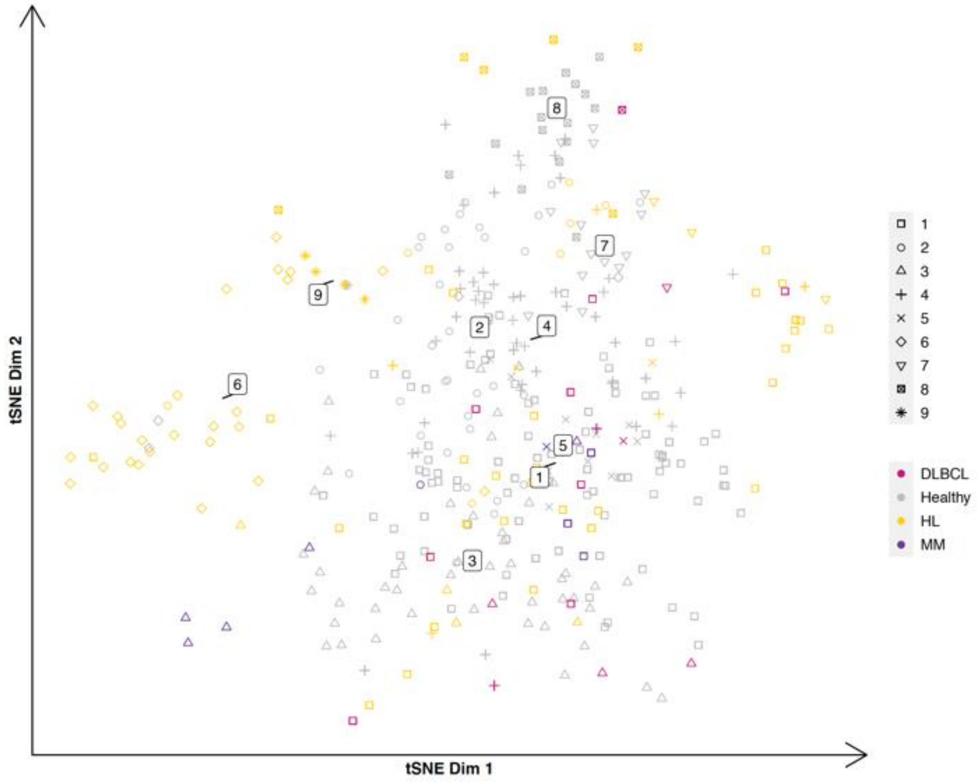


**b**

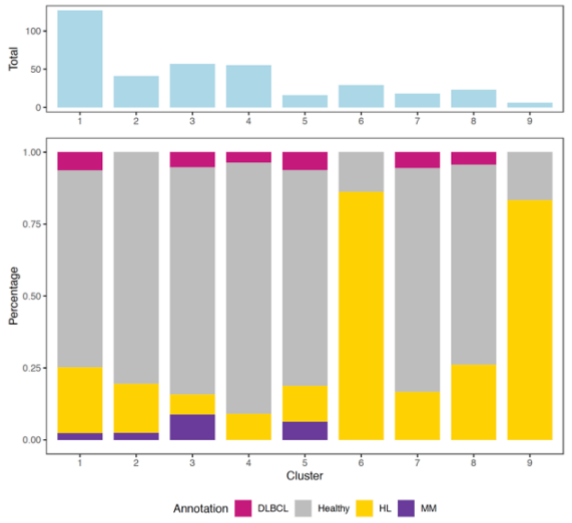


**Supp. Fig. 5.** cfDNA samples from patients with hematological cancer with estimated tumor fraction lower than 3% were used together with healthy controls for clustering analysis. **A,** tSNE visualization. **B,** cluster summarization identified by the Walktrap.

**Supplemental Figure 6. Cluster and tumor fraction distribution of misclassified samples in LOO validation for the hematological dataset.**

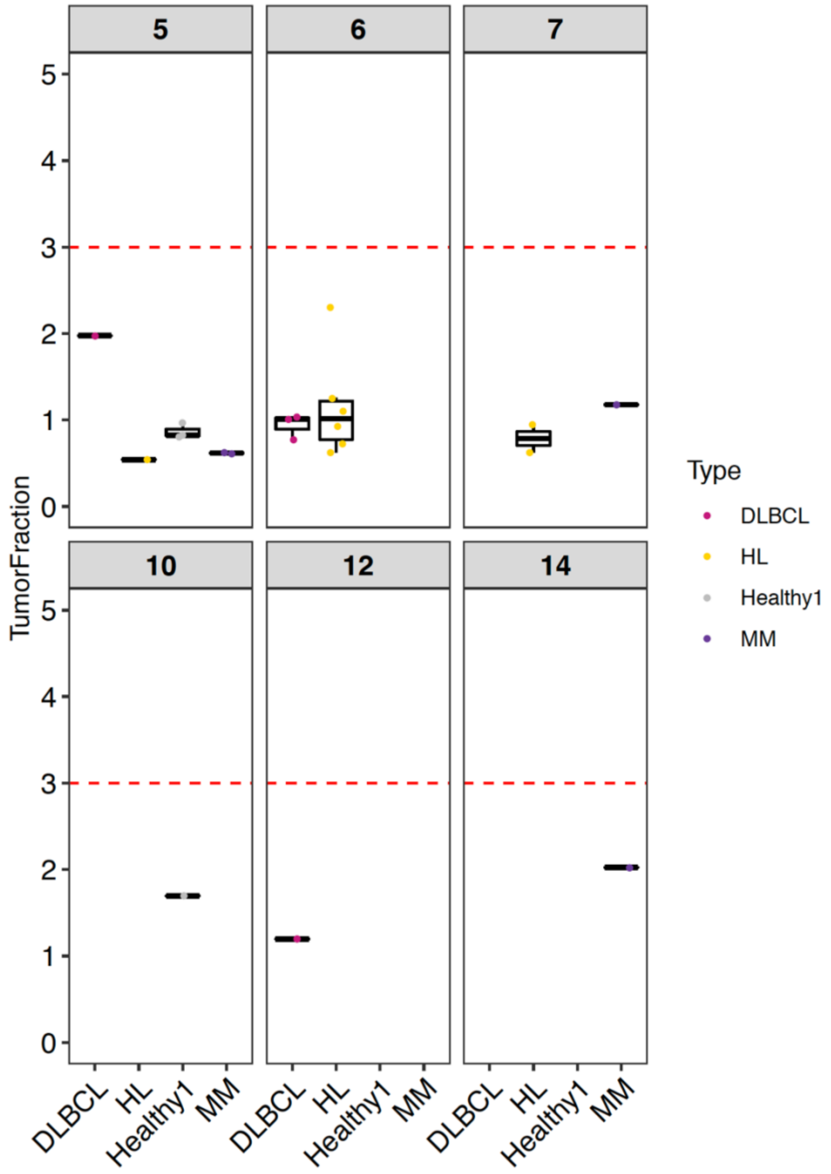


**Supp. Fig. S6.** Misclassified 18 malignant and 4 healthy control samples. Clusters that samples fell into are depicted. cfDNA samples with Hodgkin's lymphoma, (HL) are depicted in yellow, diffuse large B-cell lymphoma (DLBCL) in pink, multiple myeloma, (MM) in purple, and healthy controls are in grey.

**Supplemental Figure 7. Hematological malignancy detection with repeated 10-fold cross validation.**

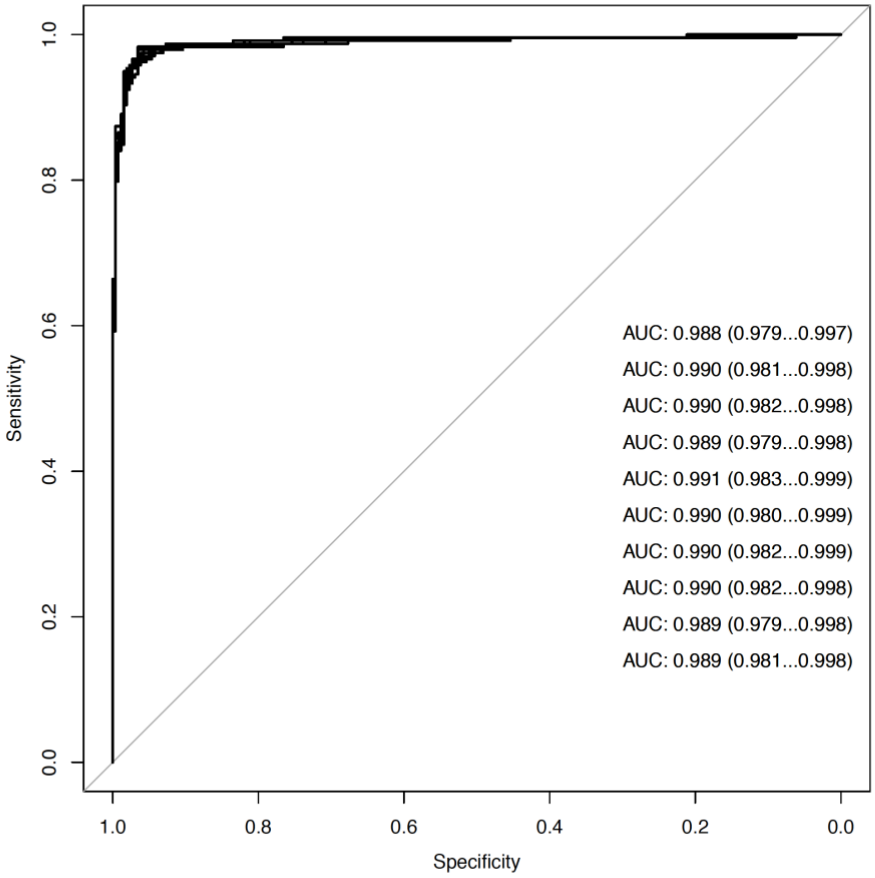


**Supp. Fig. 7.** Receiver operator characteristics for the detection of hematological malignancies using genome-wide cfDNA profiles.10-fold cross-validation was repeated for 10 times and the calculated performances of each iteration are depicted.

**Supplemental Figure 8. Clustering of the cfDNA profiles from the solid malignancy dataset.**

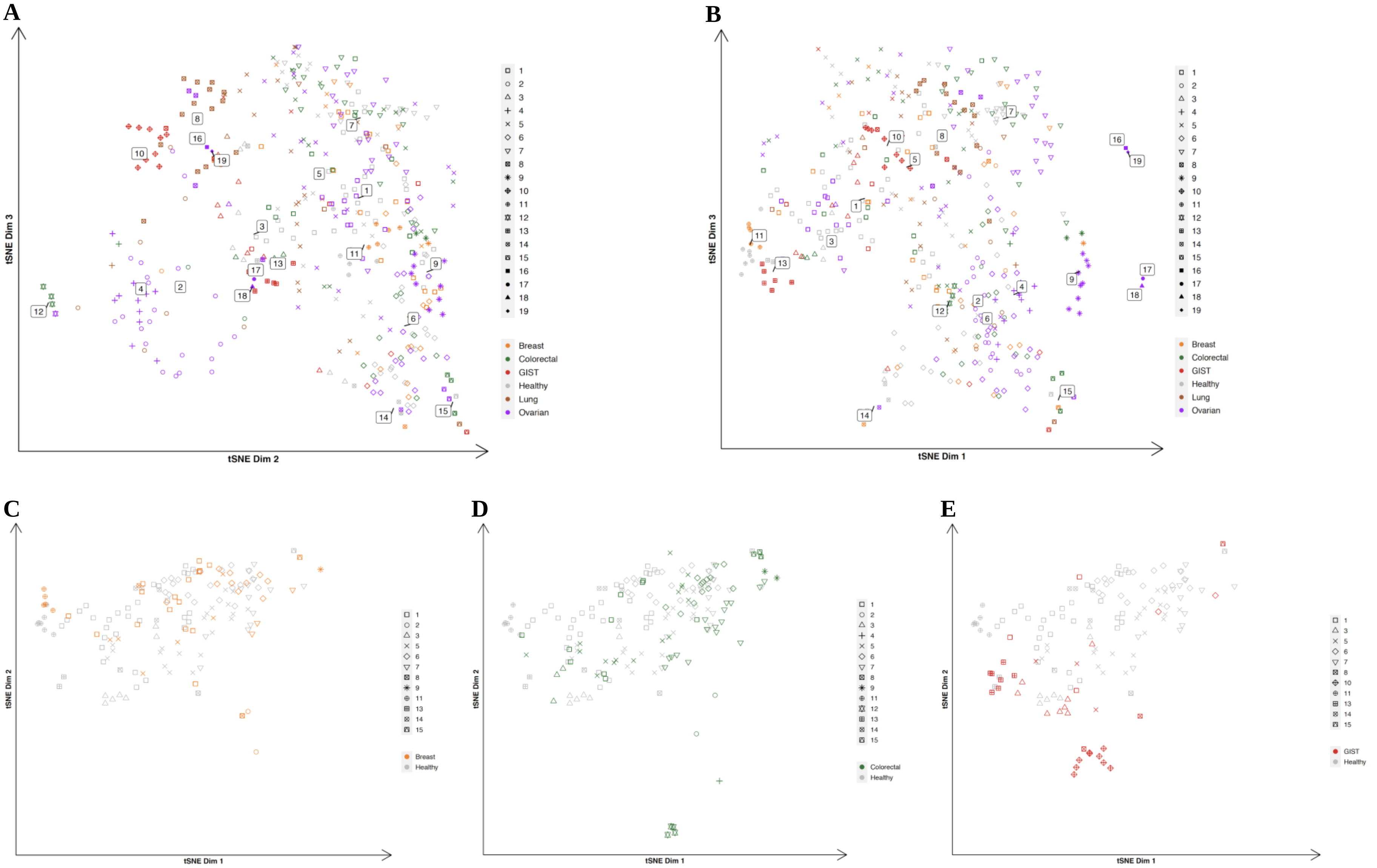

F

G

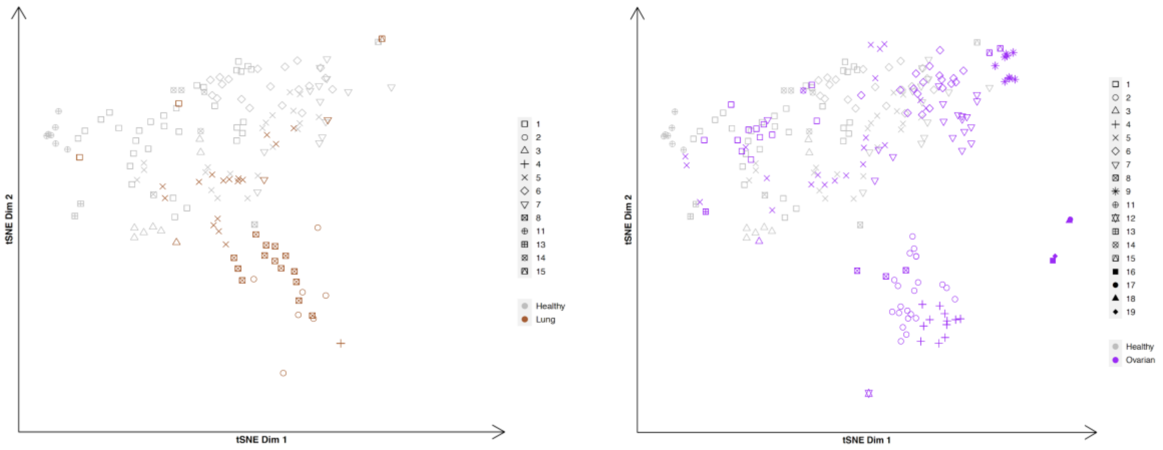


**Supp. Fig. 8.** 2D t-SNE plots for the cfDNA profiles from the solid malignancy dataset. **A,** Visualization of dimensions 2 and 3. **B,** Visualization of dimensions 1 and 3. **C-G,** cancer-specific view of breast (orange), colorectal (green), GIST (red), lung (brown), ovarian (magenta), respectively.

**Supplemental Figure 9. ichorCNA estimated tumor fraction for cfDNA samples from patients with solid malignancies.**

**A**

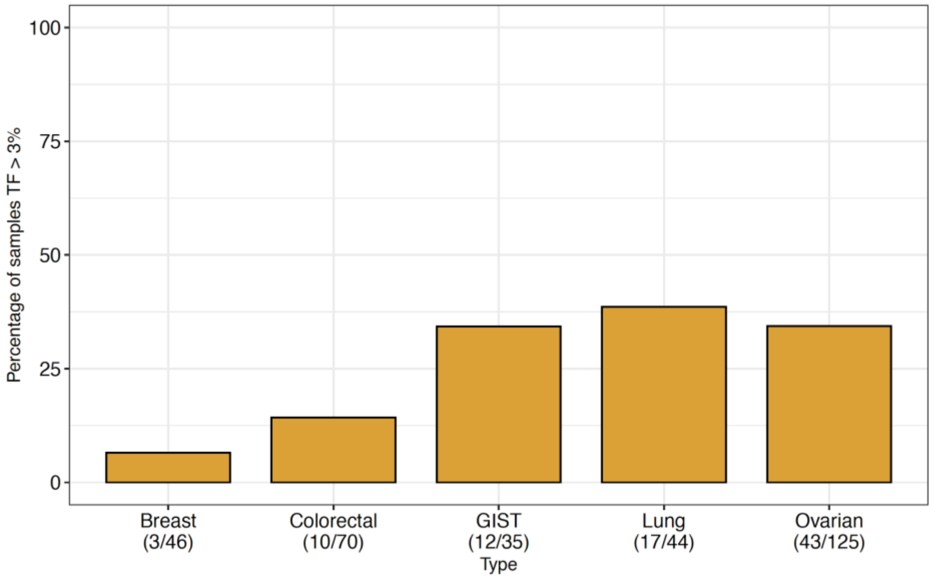


**B**

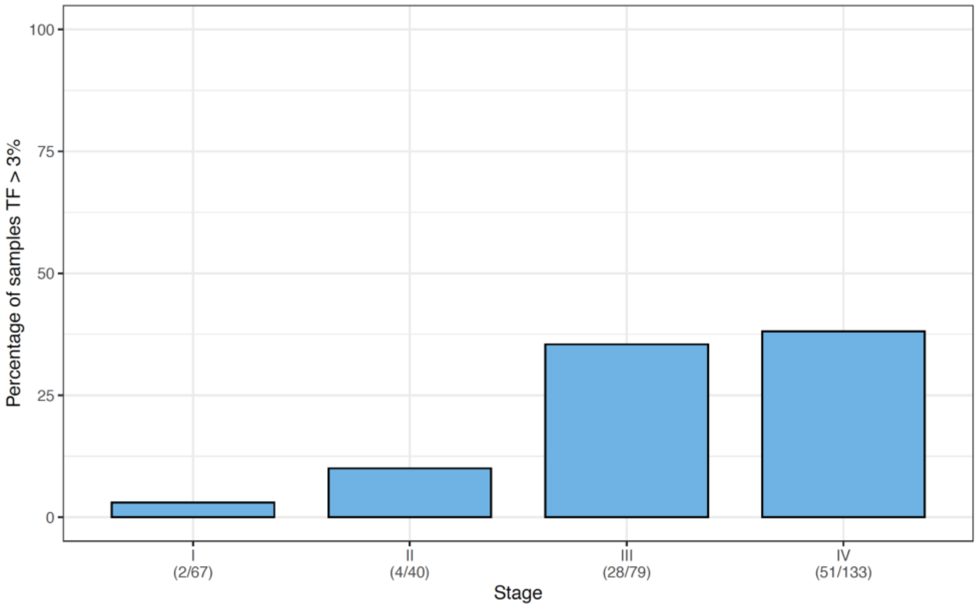


**Supp. Fig. 9. A,** Proportions of samples with tumor fraction (TF) above 3% for each type of solid cancer. **B,** Proportions of samples with tumor fraction above 3% across different stages.

**Supplemental Figure 10. Tumor fraction distribution in cfDNA samples of defined solid malignancy clusters.**

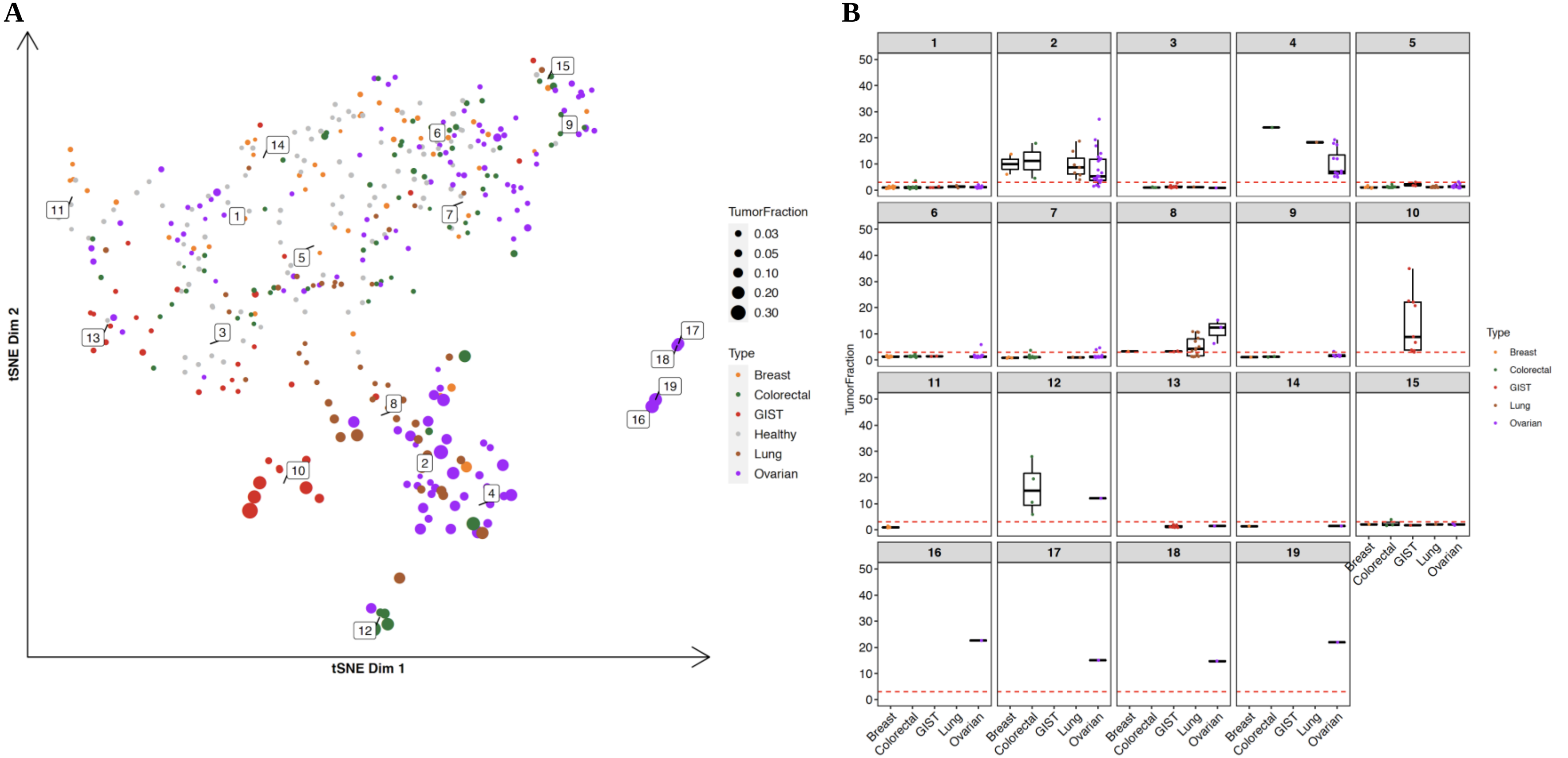


**Supp. Fig. 10. A,** cfDNA samples from solid malignancy cohort annotated with the tumor fraction and malignancy type on tSNE representation. The size of the point indicates tumor fraction levels – the larger the point, the higher the tumor fraction. **B,** Cluster-specific tumor fraction distribution for each solid cancer type.

**Supplemental Figure 11. Clustering on the log2 copy ratio profiles of solid tumor dataset.**

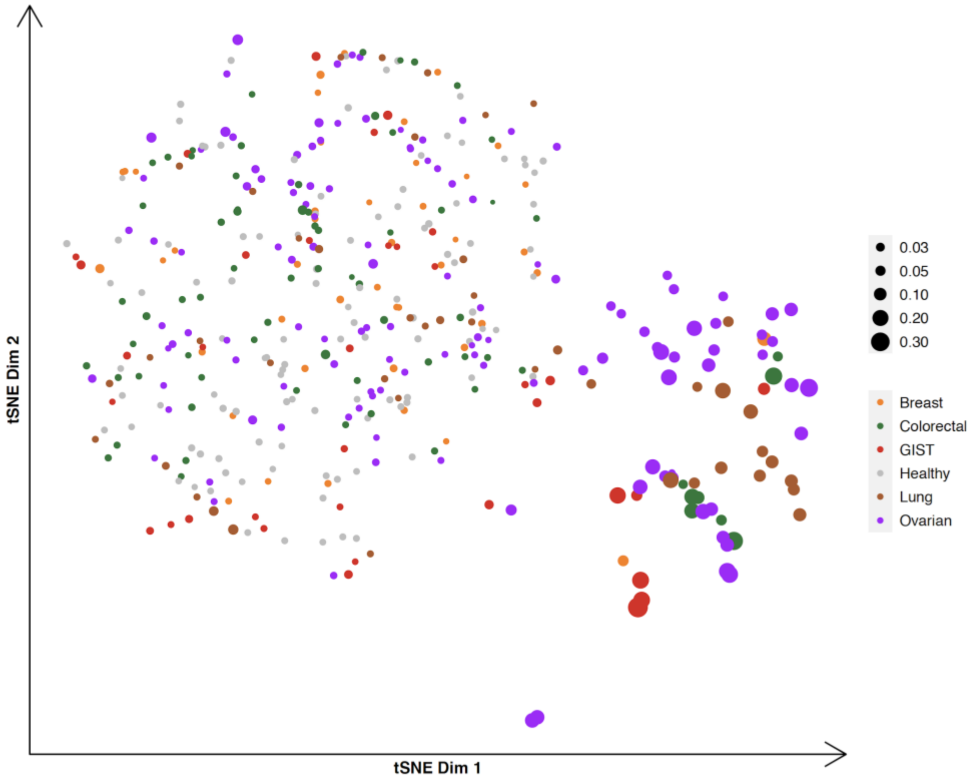


**Supp. Fig. 11.** 2D visualization of the solid tumor dataset clustering using the genome-wide log2 ratio values from ichorCNA analysis. Spearman's correlation distance-based embedding.

**Supplemental Figure 12. Clustering on cfDNA samples with low TF (<3%) from solid malignancy dataset.**

**A**

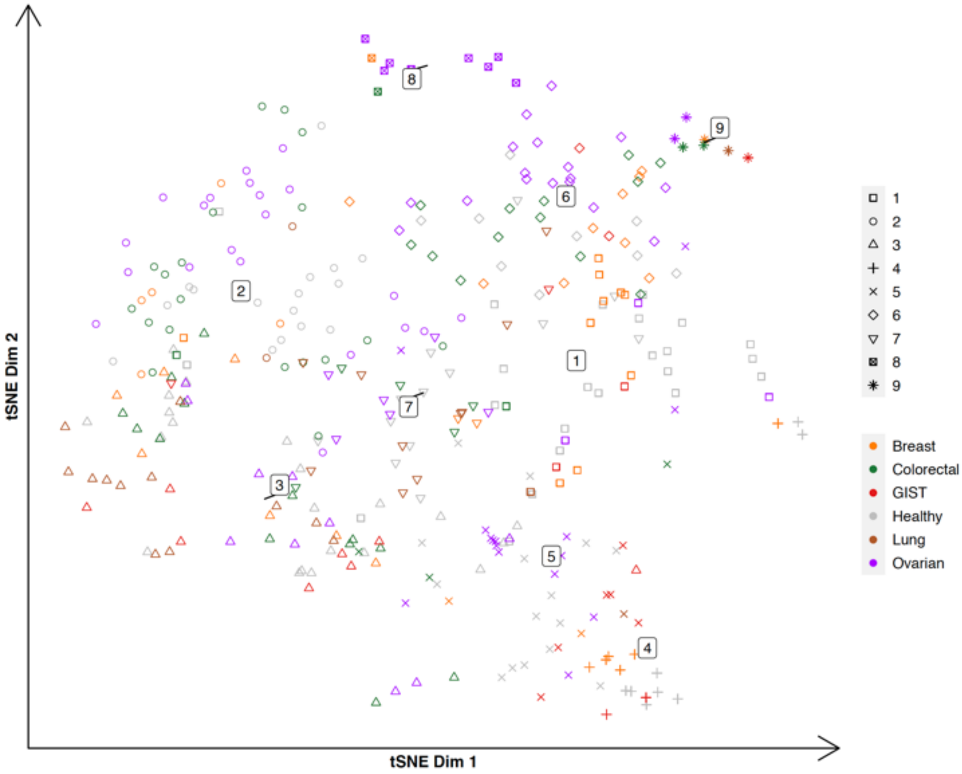


**B**

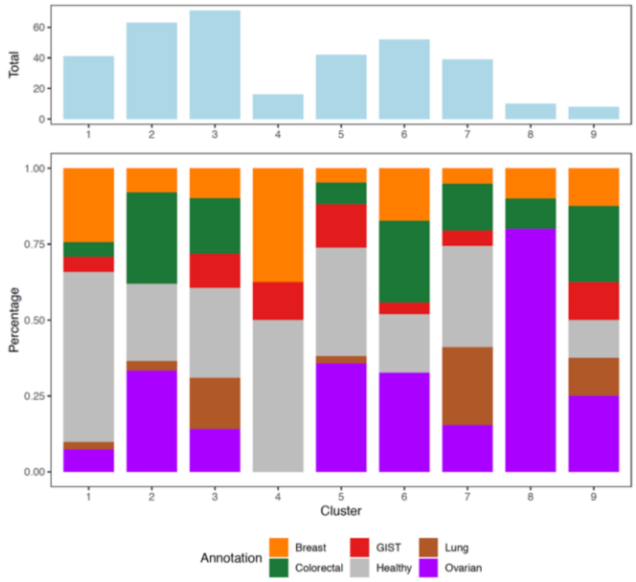


**Supp. Fig. 12.** cfDNA samples from patients with solid malignancies with estimated tumor fraction lower than 3% were used together with healthy controls for clustering analysis. **A,** tSNE visualization. **B,** cluster summarization identified by the Walktrap.

**Supplemental Figure 13. Cluster and tumor fraction distribution of misclassified samples in LOO validation for the solid tumor dataset.**

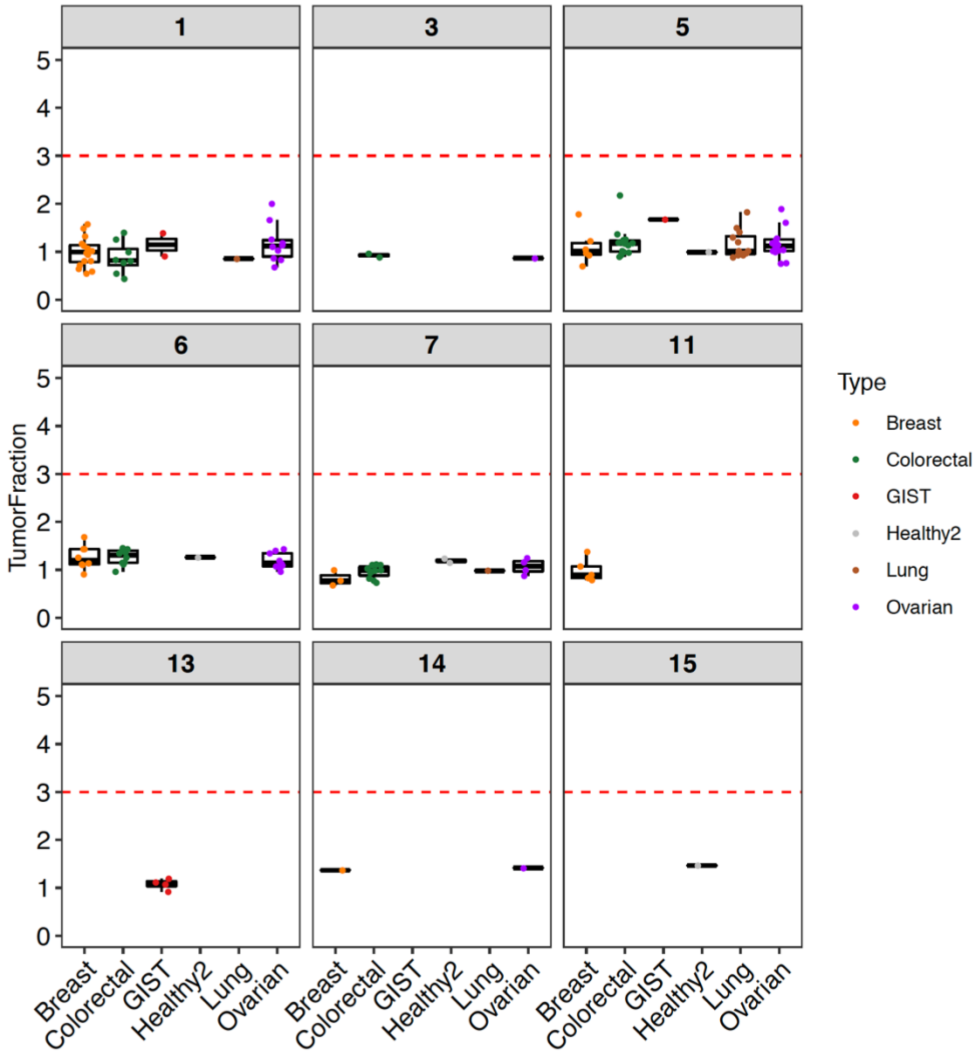


**Supp. Fig. 13.** Misclassified 143 malignant and 5 healthy control samples. Clusters that samples fell into are depicted.

**Supplemental Figure 14. Solid malignancies detection with repeated 10-fold cross validation.**

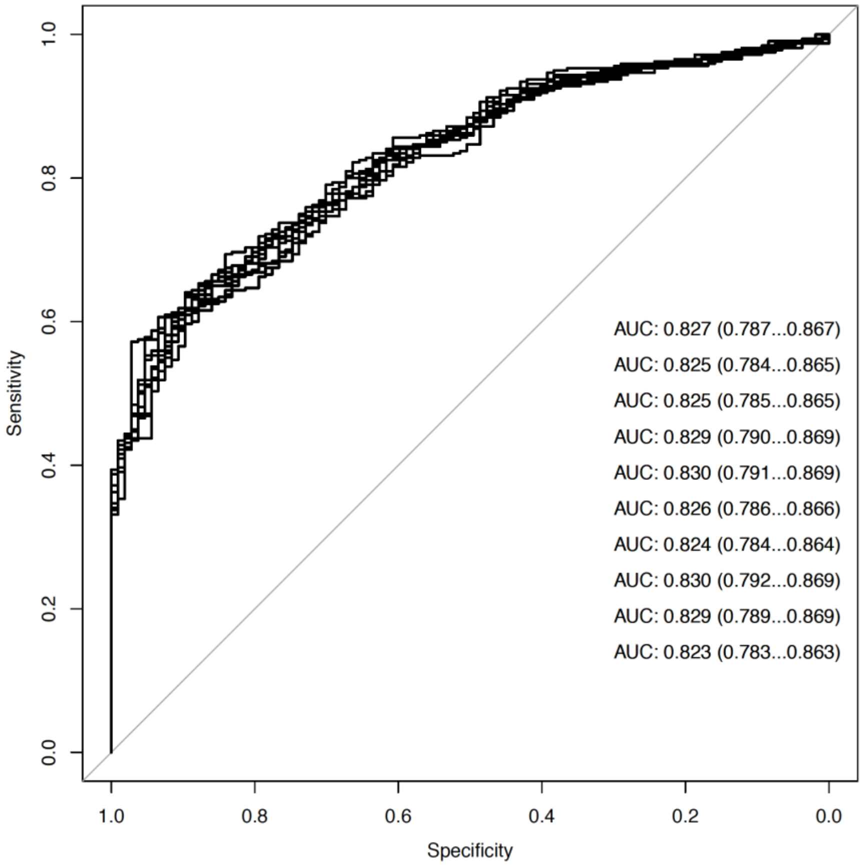


**Supp. Fig. 14.** Receiver operator characteristics for the detection of solid cancer using genome-wide cfDNA profiles.10-fold cross-validation were repeated for 10 times and the calculated performances of each iteration are depicted.

**Supplemental Figure 15**. **Annotation of primary tumor sites in samples with metastases to ovaries**.

**A**

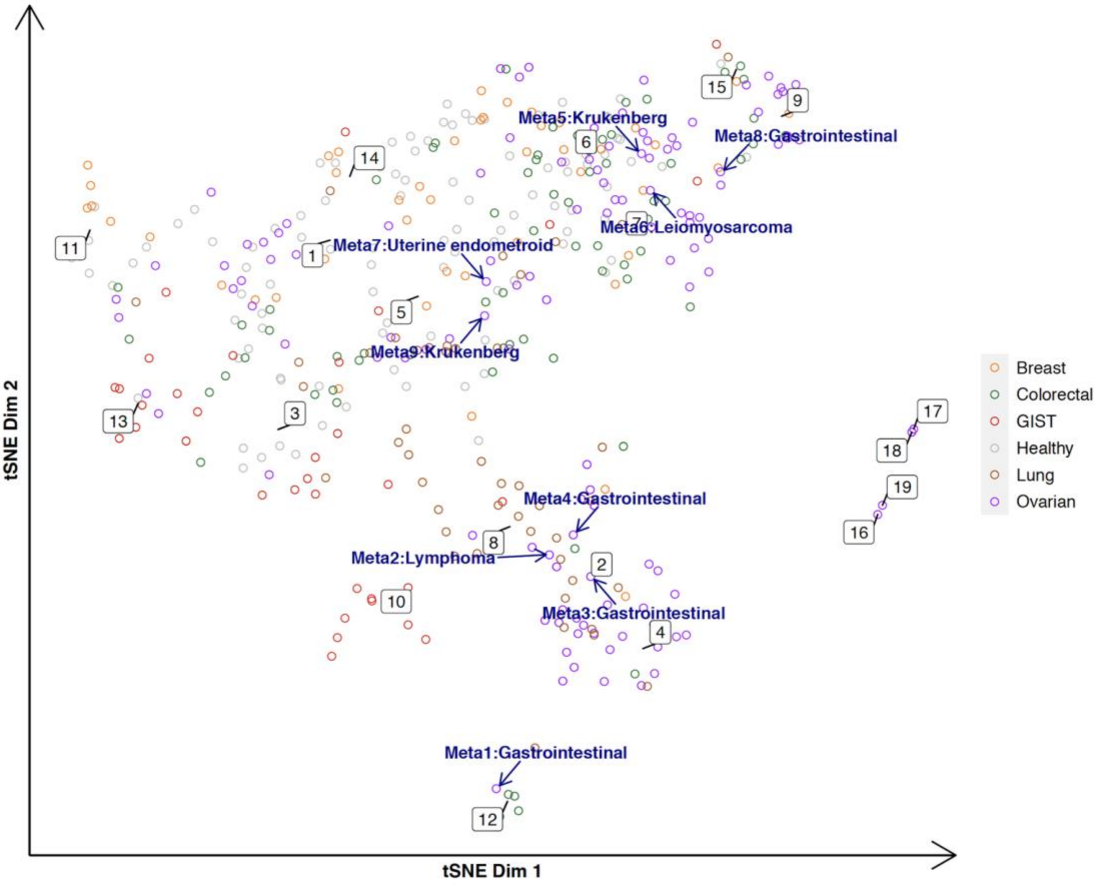


**B**

| Case | Primary site | Metastatic site | Cluster information | Classifier prediction |
|---|---|---|---|---|
| **Meta1** | Gastrointestinal | Ovarian | C12 (colocalize with colorectal) | Colorectal |
| **Meta2** | Lymphoma | Ovarian | C2 (colocalize with ovarian/intermixed) | Lung |
| **Meta3** | Gastrointestinal | Ovarian | C2 (colocalize with ovarian/intermixed) | Ovarian |
| **Meta4** | Gastrointestinal | Ovarian | C8 (colocalize with lung/intermixed) | Lung |
| **Meta5** | Krukenberg tumor (unknown primary) | Ovarian | C7 (more normal-like) | Ovarian |
| **Meta6** | Leiomyosarcoma | Ovarian | C7 (more normal-like) | Colorectal |

**Supp. Fig. 15.** Nine non-ovarian primary tumor cases with a metastasis to the ovary annotated with the primary tumor site in the t-SNE representation. **A,** Nine ovarian metastatic cases are annotated on the tSNE plot. Dark blue arrows point to the metastatic samples and the primary site is labeled. One gastrointestinal tumor sample (Meta1) with ovarian metastasis co-localized with colorectal profiles in cluster 12. Two metastatic cases (Meta3 and Meta4) fell in cluster 2, enriched for ovarian samples. The other gastrointestinal tumor case was connected to lung-like profiles in cluster 8, although the case was in close proximity with ovarian cases in the tSNE visualization. The other five metastatic cases co-localized with profiles from healthy controls. **B,** Tumor type/origin prediction.

**Supplemental Figure 16. Estimated tumor fraction in cfDNA samples from patients with benign, borderline and invasive ovarian tumors.**

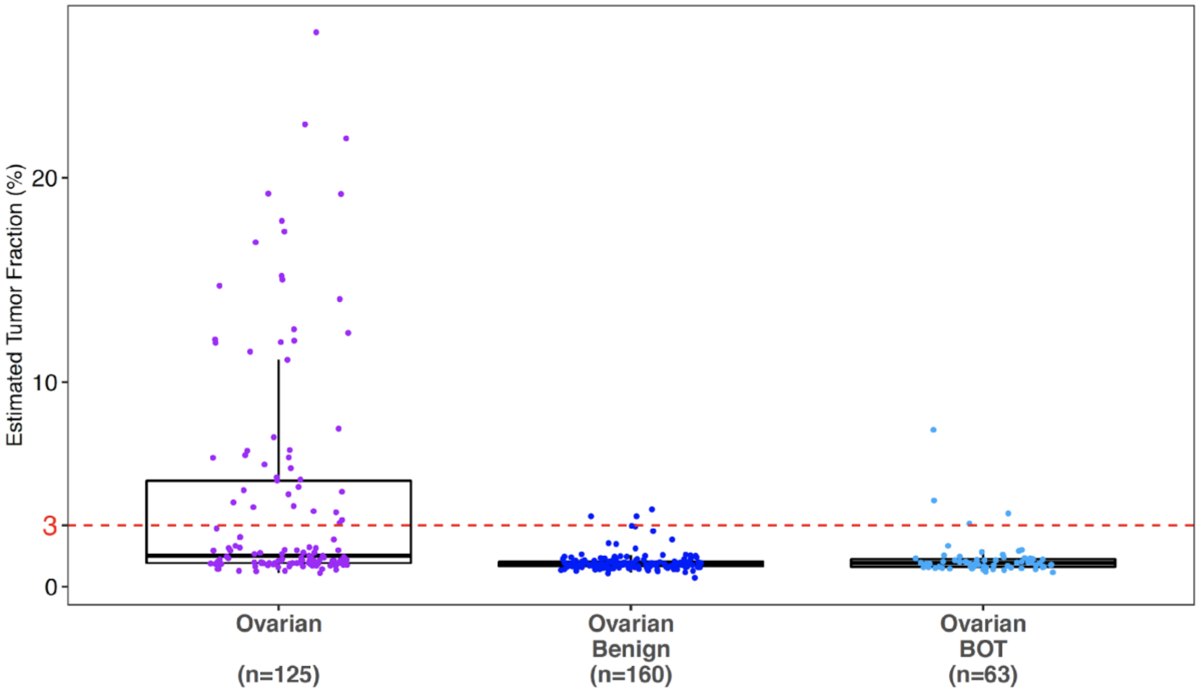


**Supp. Fig. 16.** Tumor fraction estimation for cfDNA samples from patients with ovarian tumors, including invasive (Ovarian), benign (Ovarian Benign) and borderline (Ovarian BOT).

**Supplemental Figure 17. Clustering of ovarian tumor cohort.**

**A**

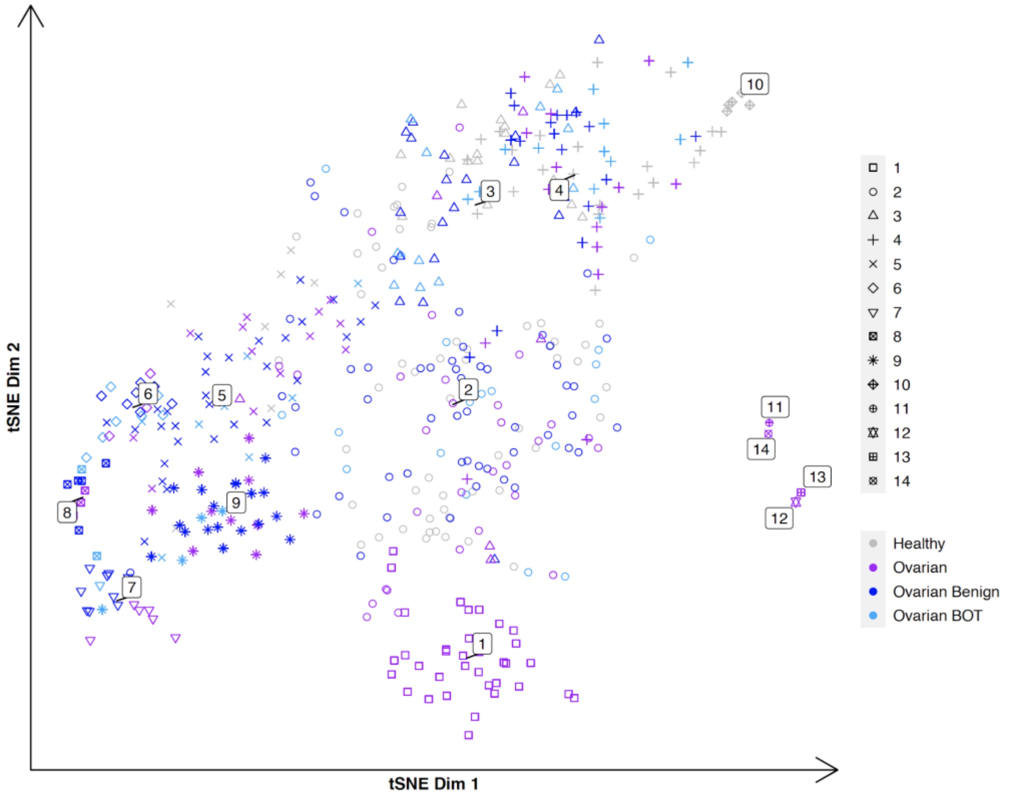


**B**

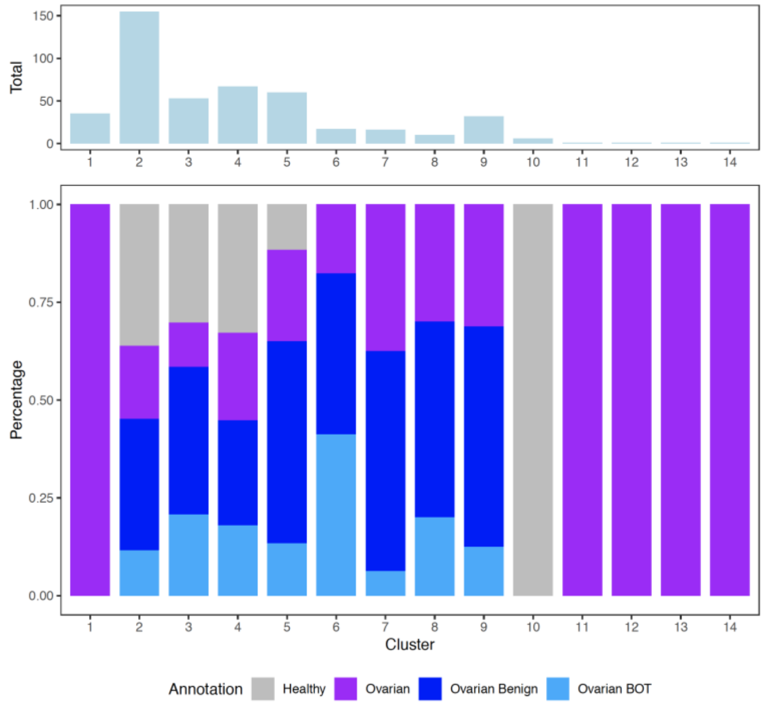


**Supp. Fig. 17. A,** tSNE visualization of ovarian tumor cohort. **B,** summary of the identified clusters.

**Supplemental Figure 18. ROC curve for discrimination between ovarian benign and malignant cases.**

**A**

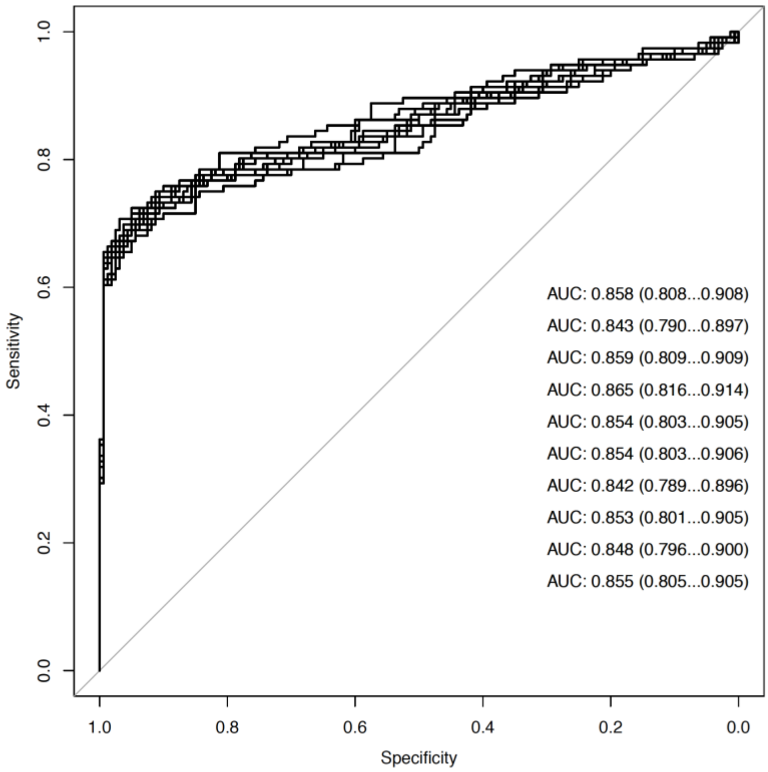


**B**

| | | Prediction | | |
|---|---|---|---|---|
| | | Benign | Invasive | Total |
| True | Benign | 159 | 1 | 160 |
| | Invasive | 47 | 69 | 116 |
| Accuracy | | | | 0.8261 |
| Sensitivity | | | | 0.5948 |
| Specificity | | | | 0.9938 |
| PPV | | | | 0.9857 |
| NPV | | | | 0.7718 |
| Balanced Accuracy | | | | 0.7943 |

**Supp. Fig. 18.** 160 ovarian benign and 116 invasive samples were analyzed. **A,** ROC curves of 10-fold cross validation repeated for 10 times are shown. **B,** Prediction results from LOO analysis.

**Supplemental Figure 19. ROC curve for discrimination between ovarian benign and borderline + malignant cases.**

**A**

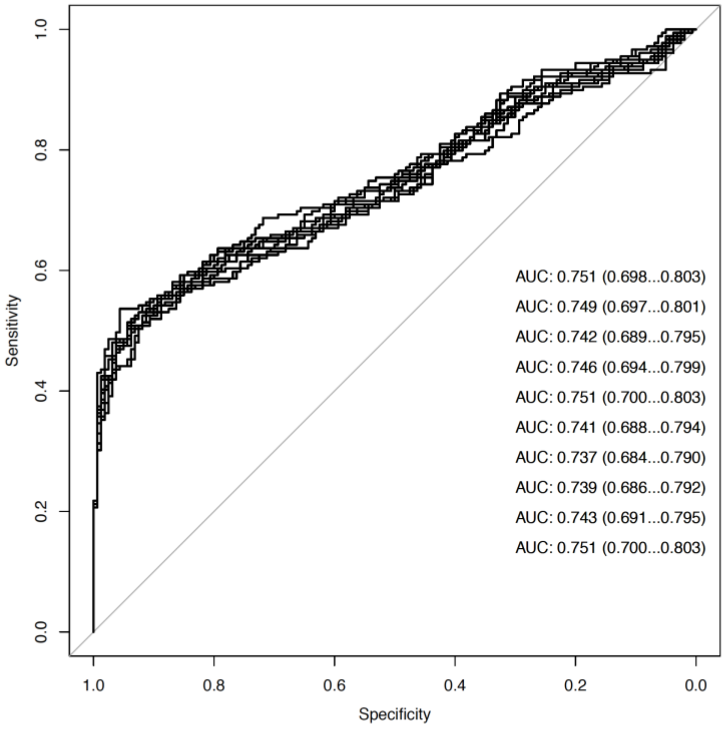


**B**

| | | Prediction | | |
|---|---|---|---|---|
| | | Benign | BOT/Invasive | Total |
| True | Benign | 154 | 6 | 160 |
| | BOT/Invasive | 88 | 91 | 179 |

| | |
|---|---|
| Accuracy | 0.7227 |
| Sensitivity | 0.5084 |
| Specificity | 0.9625 |
| PPV | 0.9381 |
| NPV | 0.6364 |
| Balanced Accuracy | 0.7354 |

**Supp. Fig. 19.** 160 ovarian benign and 179 borderline + invasive samples were analyzed. **A,** ROC curves of 10-fold cross validation repeated for 10 times are shown. **B,** Prediction results from LOO analysis.

**Supplemental Figure 20. Cumulative variance in principal components analysis.**

**A** **B**

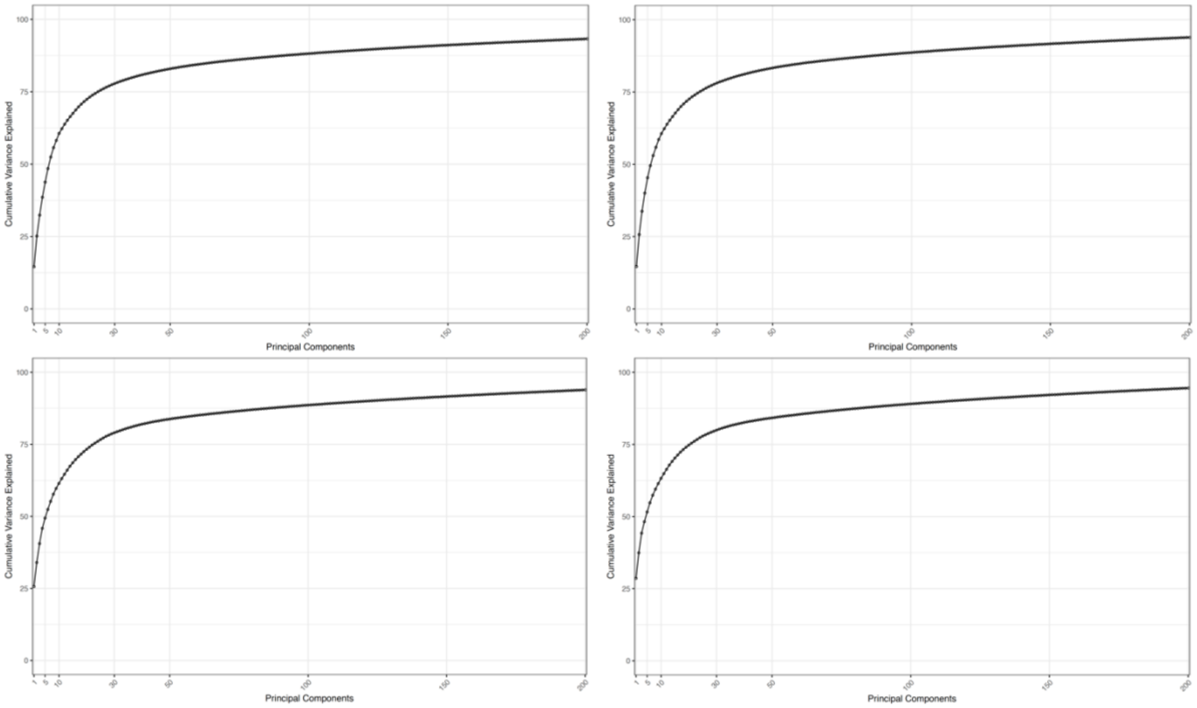


**Supp. Fig. 20.** Cumulative variance scree plot. **A,** Cumulative variance explained by the first 200 components in the dataset for hematological (n=498) and solid malignant dataset (n=427), respectively. **B,** Cumulative variance explained by the first 200 components using 90% randomly downsampled data in hematological and solid tumor dataset, respectively.

**Supplemental Figure 21. Non-trivial principal components.**

**A**

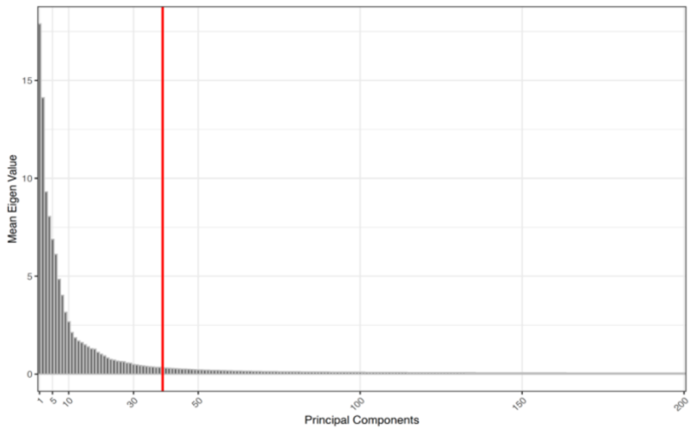


**B**

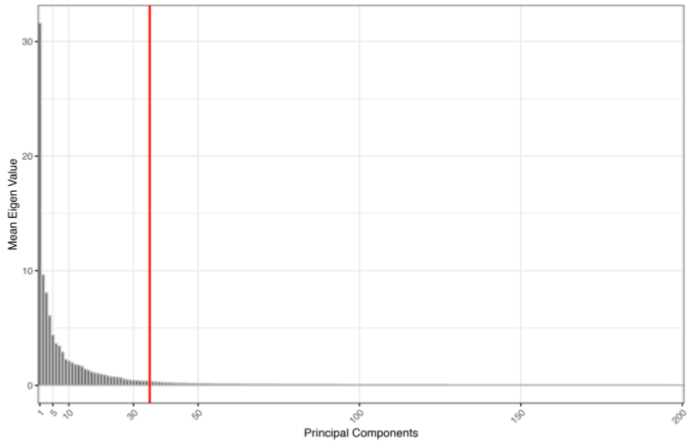


**Supp. Fig. 21.** Bar plot showing averaged eigen values across five replicas and the high signal thresholds (red vertical line) for the number of principal components. **A,** The permutation test in 90% of the hematological dataset and all five replicates resulted in a threshold of 39. **B,** The permutation test in 90% of the solid malignant dataset and all five replicates resulted in a threshold of 35.

**Supplemental Figure 22. Pairwise distance in the hematological dataset.**

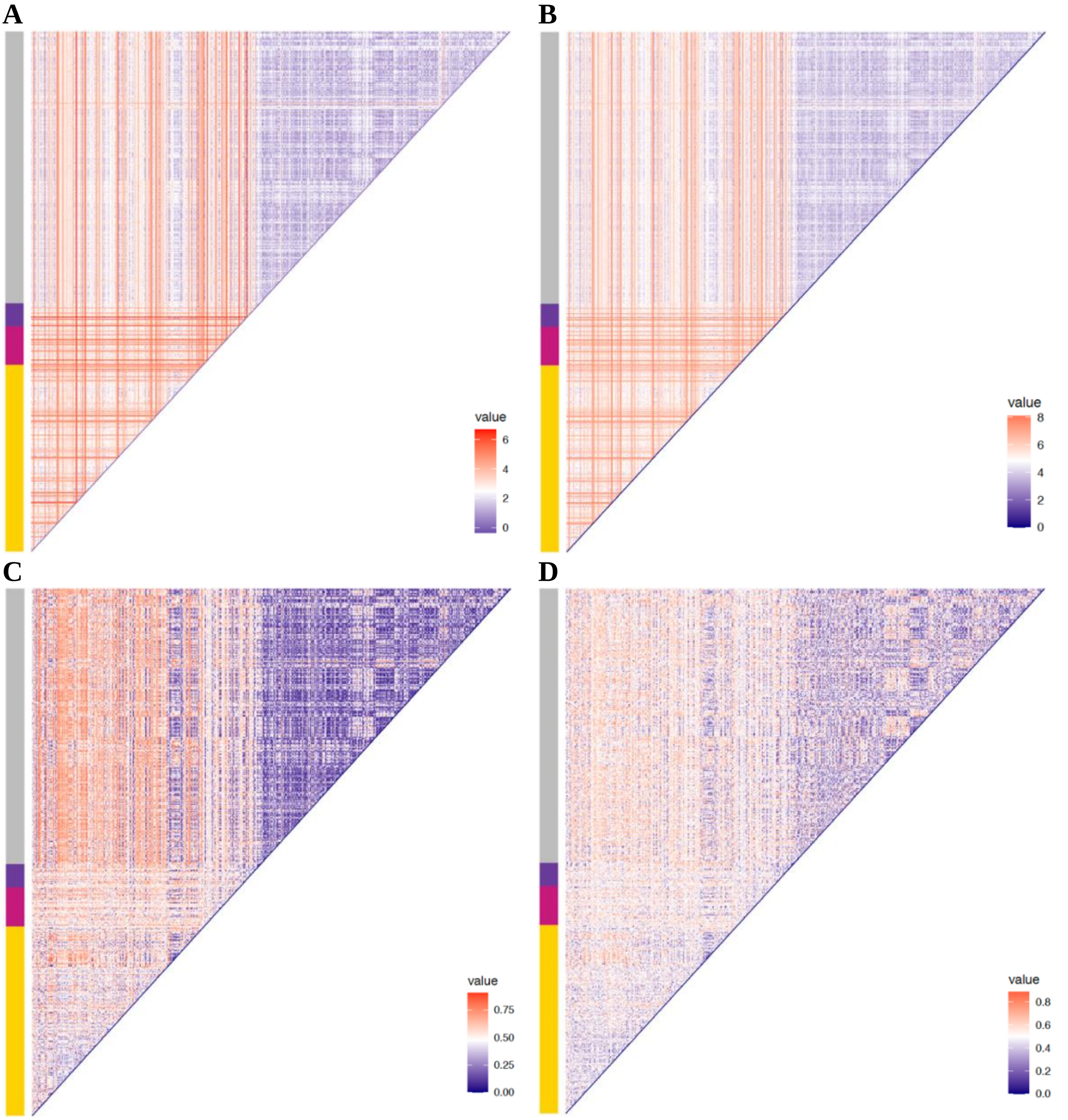


**Supp. Fig. 22.** Heatmap showing the pairwise distance of 498 samples in the hematological dataset. For better visualization, the Euclidean and Manhattan distances were transformed with log2 scale. The side vertical bar indicates the type of samples. The color code is identical to **Figure 2B**. **A,** Euclidean distance. **B,** Manhattan distance. **C,** Pearson correlation distance (d = (1-*pearson correlation*)/2). **D,** Spearman's correlation distance (d = (1-*spearman's correlation*)/2).

**Supplemental Figure 23. Pairwise distance in the solid tumor dataset.**

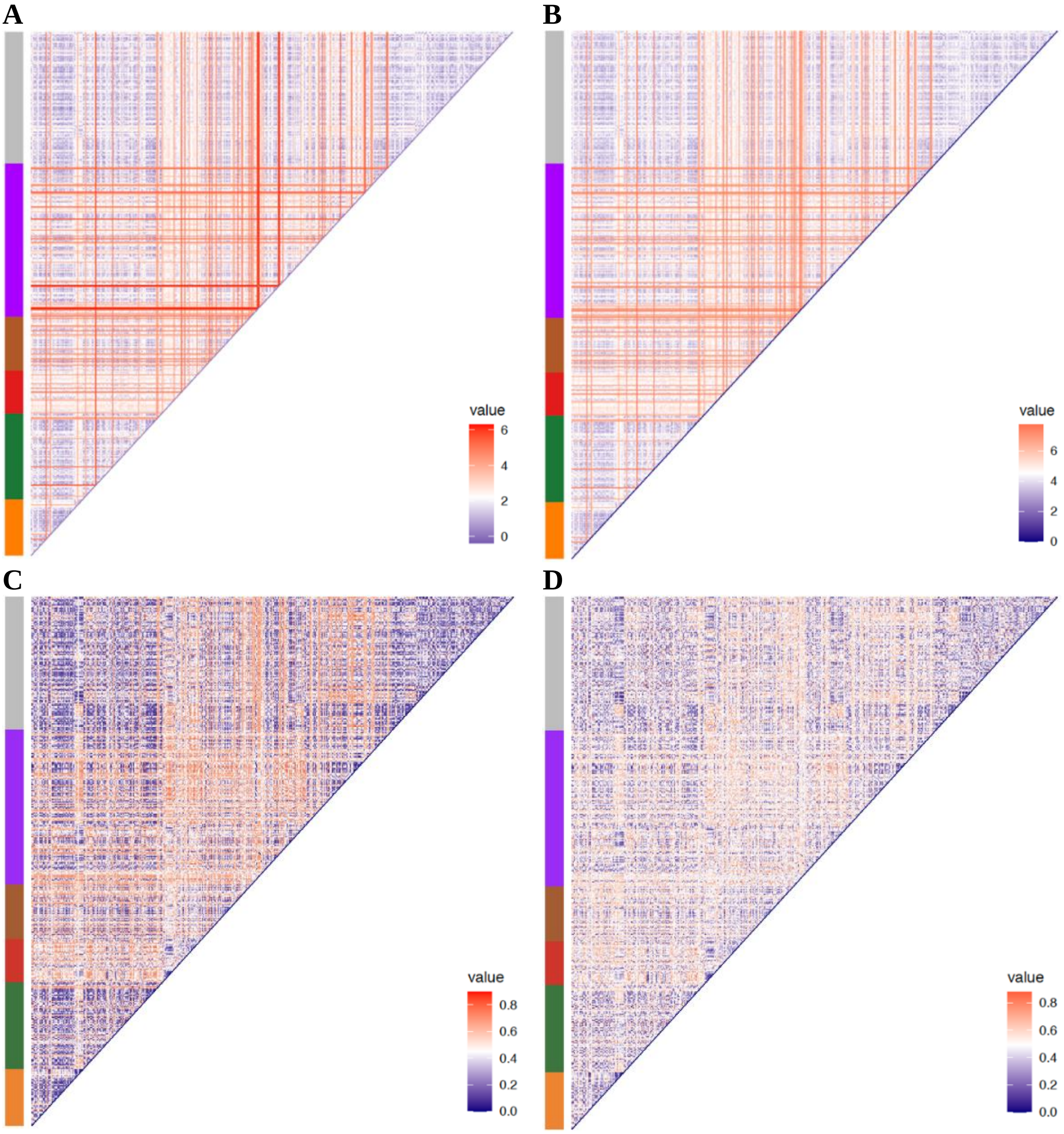


**Supp. Fig. 23.** Heatmap showing the pairwise distance of 427 samples in the solid malignant dataset. For better visualization, the Euclidean and Manhattan distances were transformed with log2 scale. The side vertical bar indicates the type of samples. The color code is identical to **Figure 4B**. **A,** Euclidean distance. **B,** Manhattan distance. **C,** Pearson correlation distance (d = (1-*pearson correlation*)/2). **D,** Spearman's correlation distance (d = (1-*spearman's correlation*)/2).

**Supplemental Figure 24. Walktrap community modularity for the hematological dataset.**

**A**

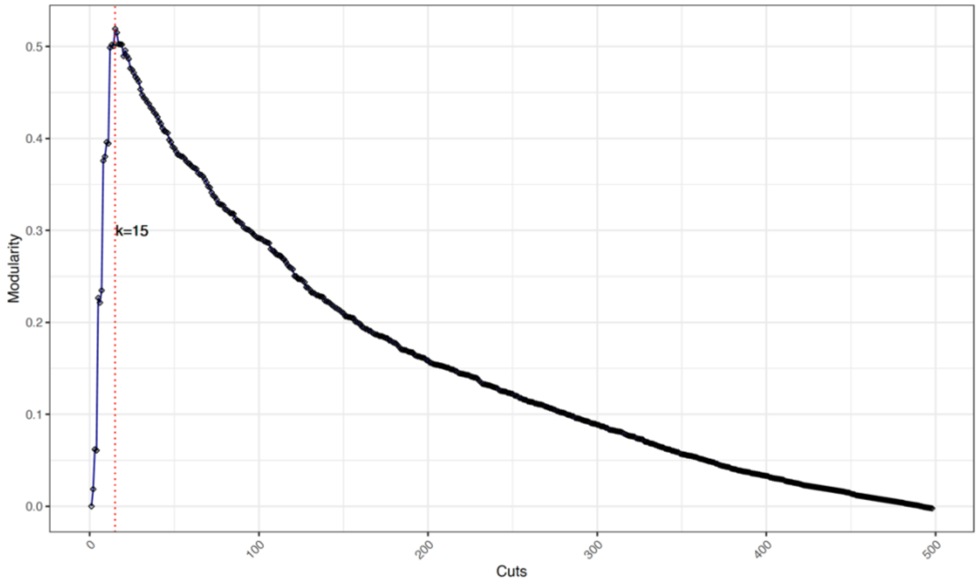


**B**

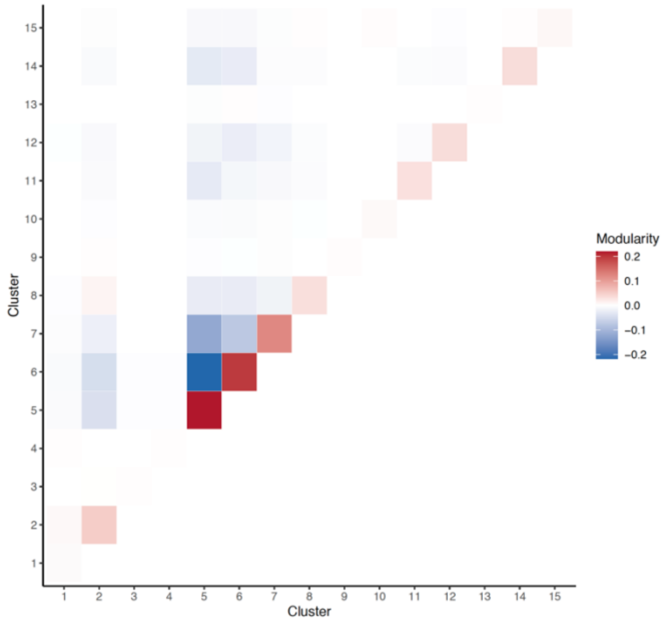


**Supp. Fig. 24. A,** The modularity score at different number of community cutoffs using the Walktrap community detection. The cutoff with the highest modularity score is indicated in red vertical line. **B,** modularity score (on log2 scale) of pairwise clusters defined by the optimal number of communities from **A.** High modularity score indicates dense connection between nodes within the communities but sparse connections between nodes in different communities**.**

**Supplemental Figure 25. Walktrap community modularity for the solid tumor dataset.**

**A**

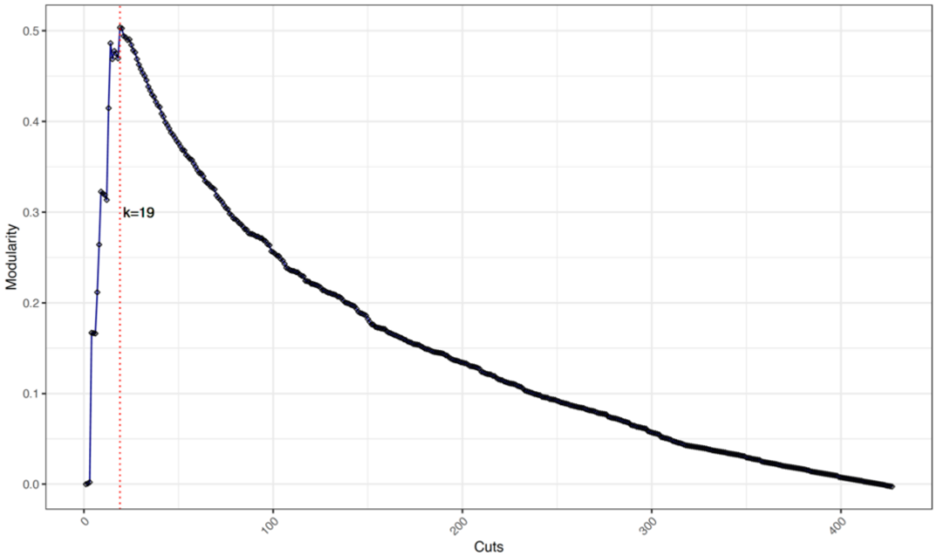


**B**

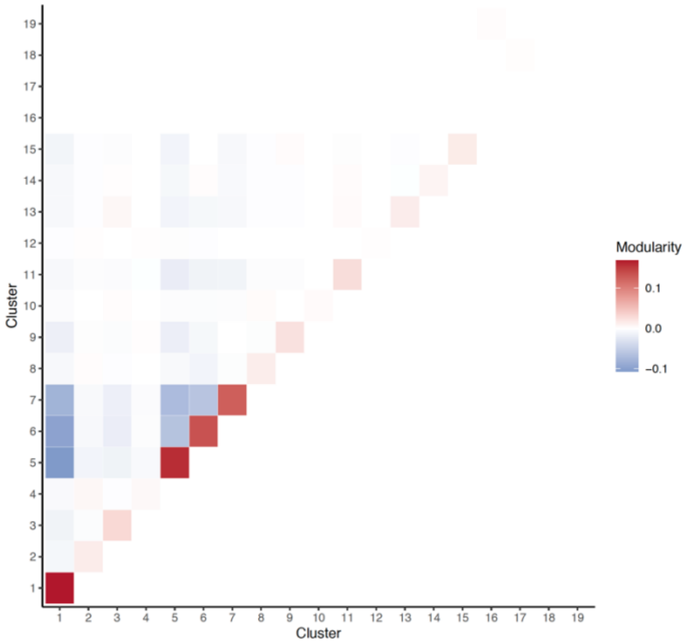


**Supp. Fig. 25. A,** The modularity score at different number of community cutoffs using the Walktrap community detection. The cutoff with the highest modularity score is indicated in red vertical line. **B,** modularity score (on log2 scale) of pairwise clusters defined by the optimal number of communities from **A.** High modularity score indicates dense connection between nodes within the communities but sparse connections between nodes in different communities**.**

**Supplemental Figure 26. ROC for cancer profile prediction using original genome-wide features.**

**A**

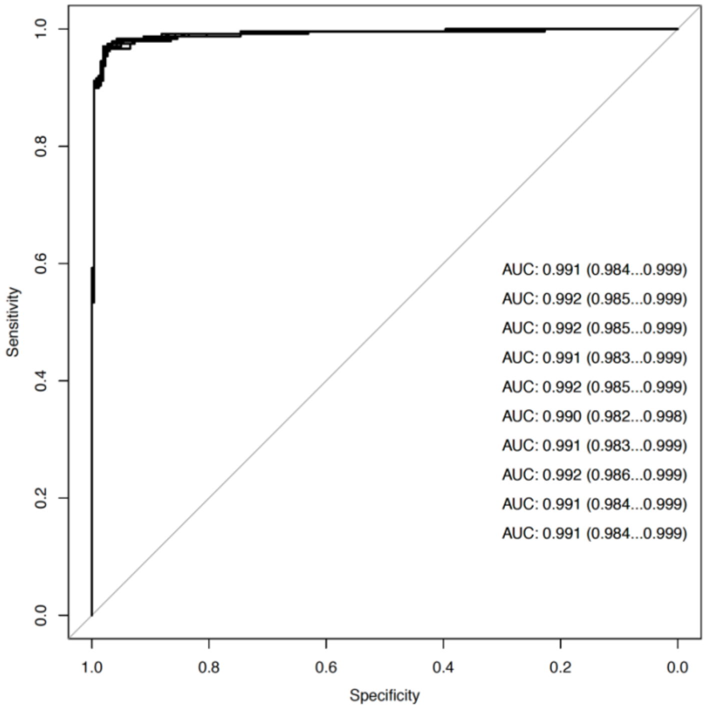


**B**

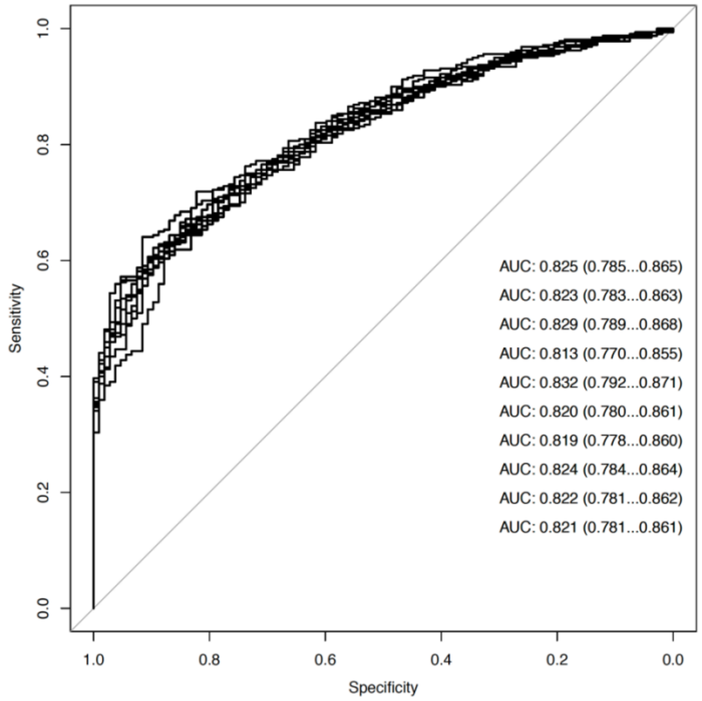


**Supp. Fig. 26. A,** Ten replicates of ROC analysis of 10-fold CV for identifying hematological cancer profiles using the original genome-wide features. **B,** ROC analysis of 10-fold CV for identifying solid malignant profiles using the original genome-wide features.

**Supplemental Figure 27. Evaluation of possible confounding factors using clustering.**

A

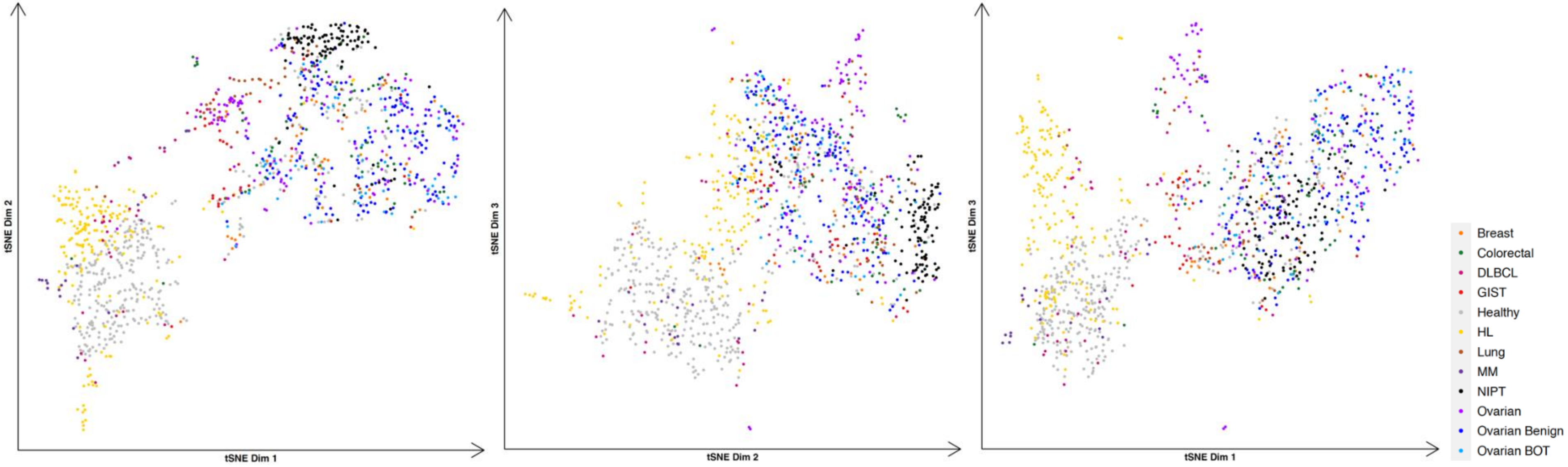


B

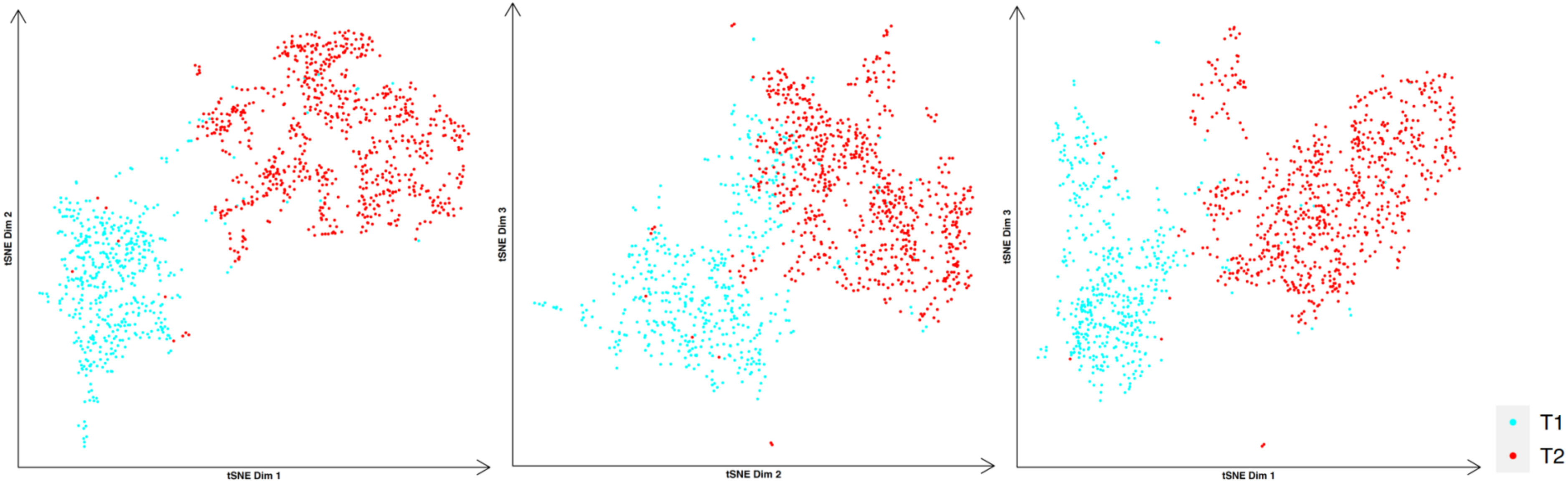

**Supplemental Figure 27. Continued**

**C**

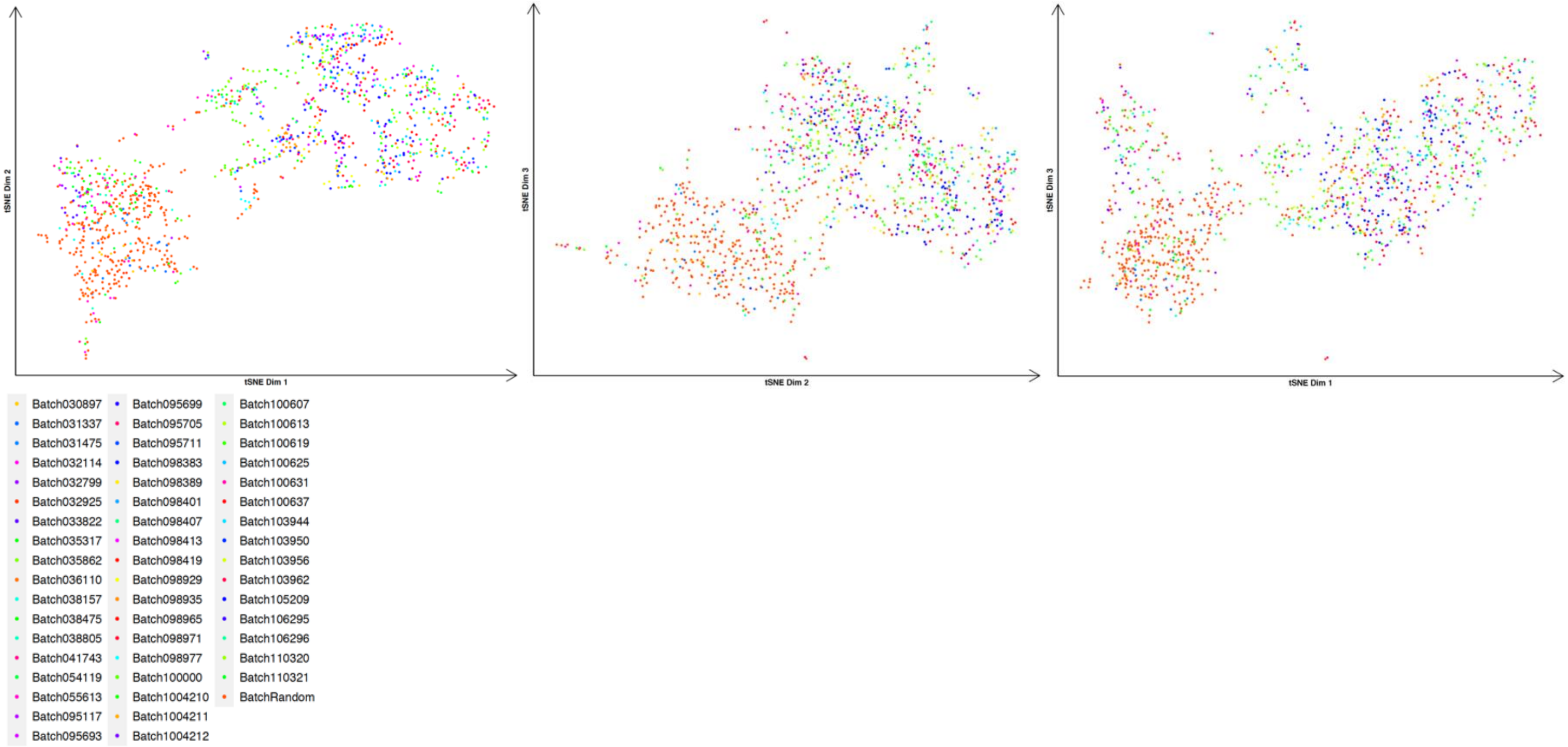


**D**

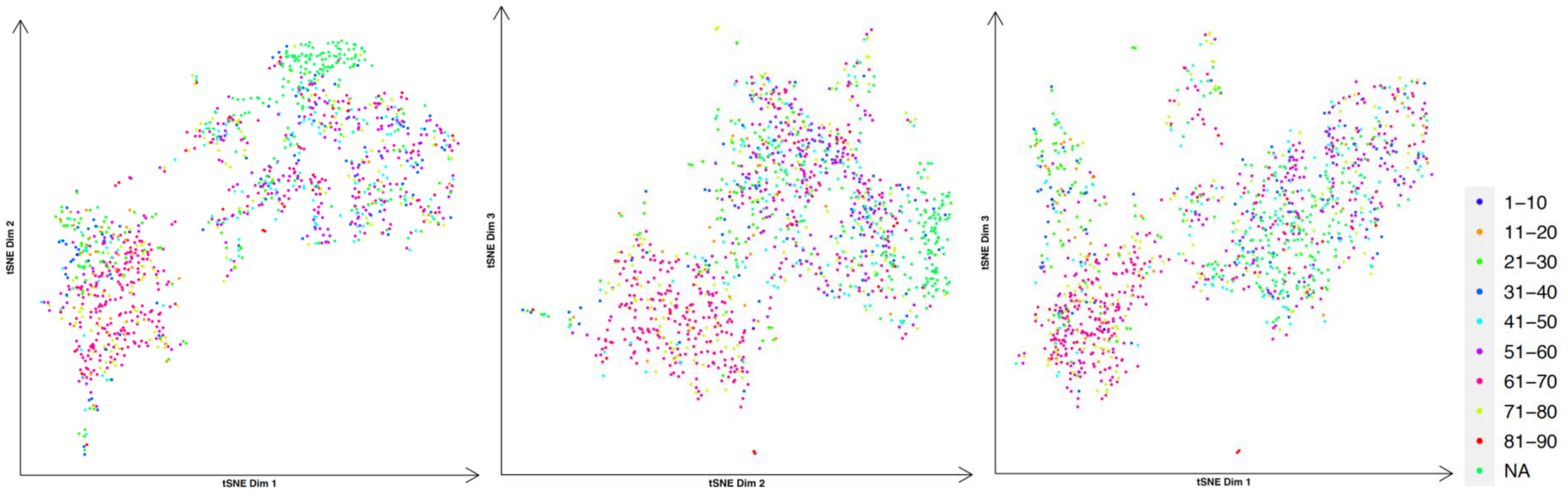
tSNE Dim 1
tSNE Dim 2
tSNE Dim 3
1–10
11–20
21–30
31–40
41–50
51–60
61–70
71–80
81–90
NA

E

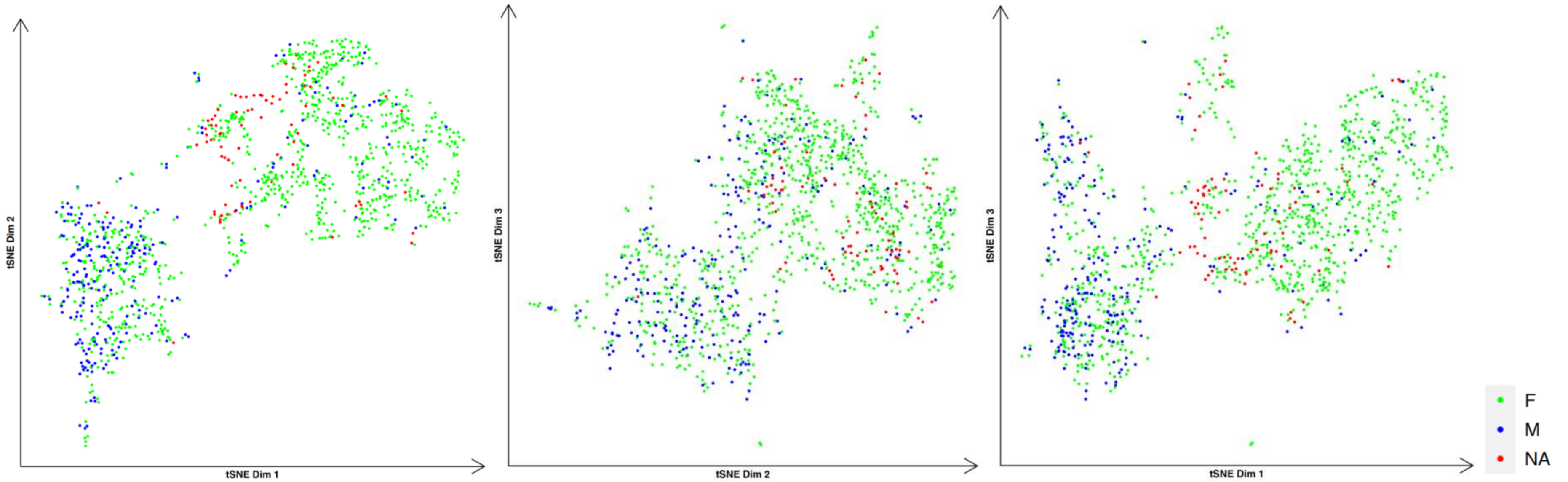
tSNE Dim 1
tSNE Dim 2
tSNE Dim 3
F
M
NA

**Supp. Fig. 27.** tSNE visualization on the entire dataset, including the hematological tumor and solid tumor. In addition, we included 102 pregnant cfDNA profiles. **A,** tSNE visualization annotated by sample type. **B,** tSNE visualization annotated by library preparation method. The separated distribution of samples, based on the preparation kits (T1 and T2), indicates the bias could confound the analysis. **C,** annotation by sequencing batch. If no less than 10 samples were pooled together and processed in one sequencing batch, the batch is labeled with that specific run. The rest of the samples that were processed in separate runs, are labeled with a random batch. **D,** annotation by age group. **E,** annotation by sex. 'F' represents female, 'M' represents male and 'NA' for missing information.

**Supplemental Figure 28. Genomic profiles of multiple samples from different library preparation kits.**

**A**

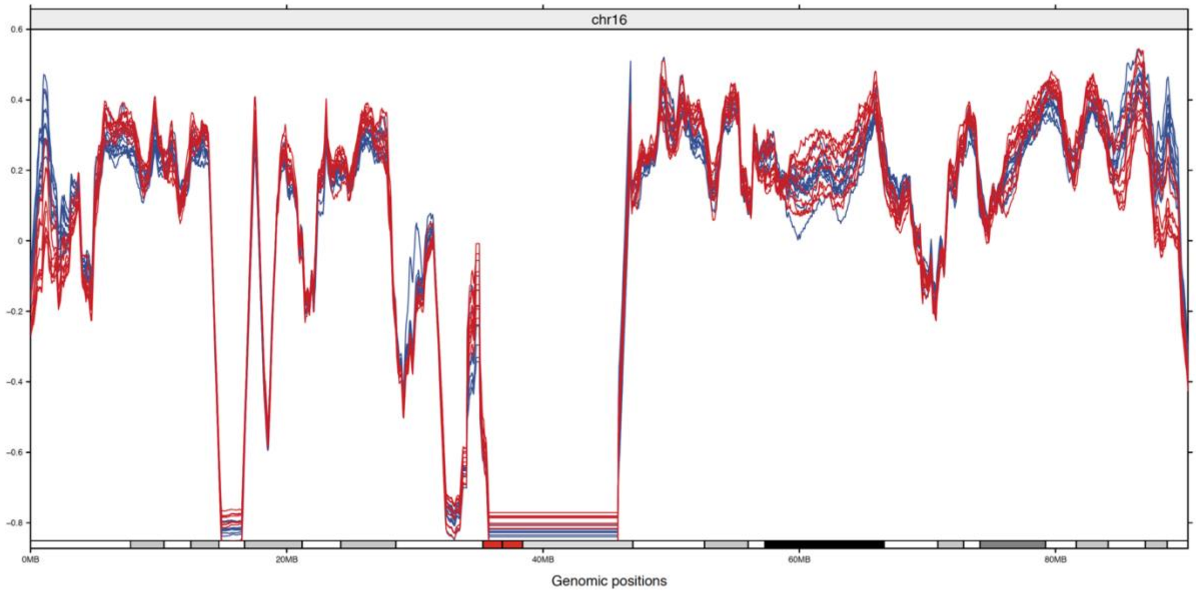


**B**

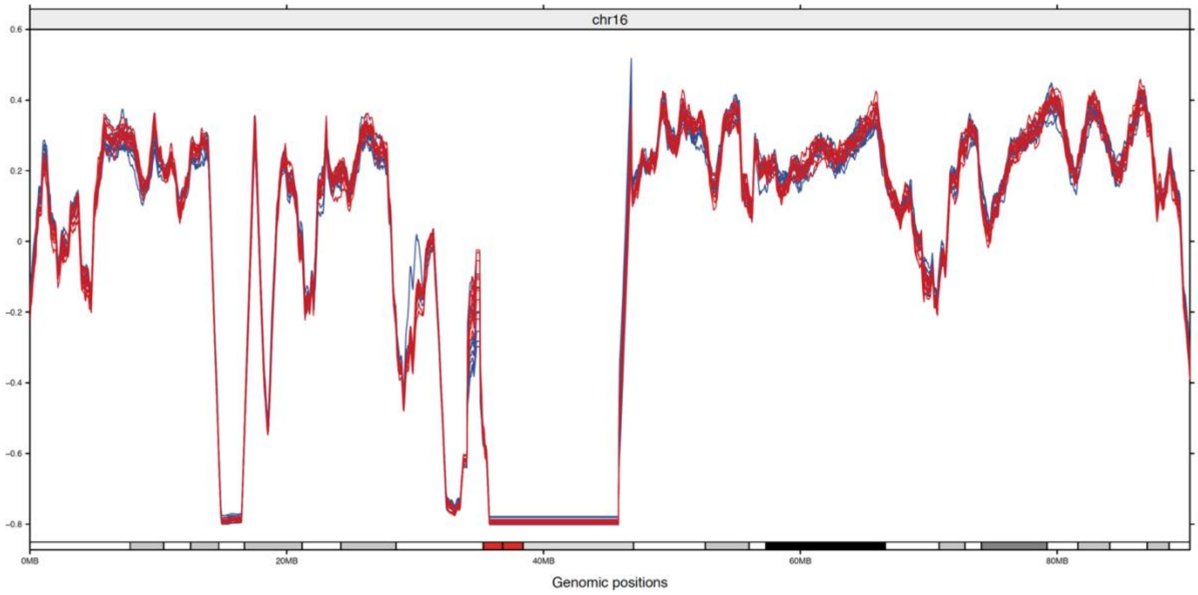


**Supp. Fig. 28.** Smoothed coverage profiles of 10 cfDNA samples prepared using KAPA HyperPrep (color blue) and 10 samples using the TruSeq ChIP (color red) library preparation kits without GC correction **A**, and with GC correction **B** mapped to chromosome 16.

**Supplemental Figure 29. Tumor fraction estimation comparison between 2 reference panels.**

**A**

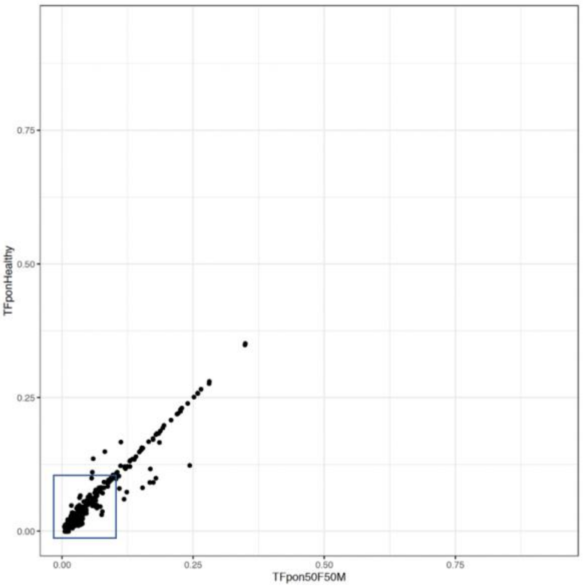


**B**

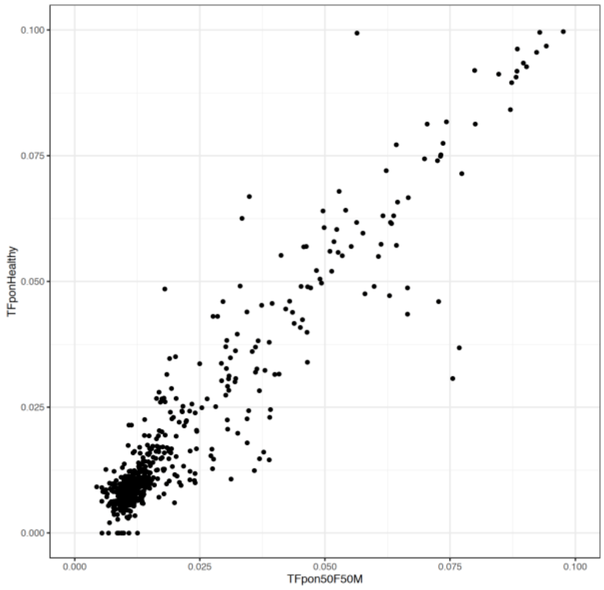


**Supp. Fig. 29.** To investigate the presence of potential technical bias resulting from library preparation methods on the tumor fraction estimation, we built two panels of normal, in which one panel was constructed from 100 samples from the TruSeq ChIP and the other panel from 100 samples prepared using the KAPA HyperPrep library preparation kit. A total of 654 tumor samples either processed with TruSeq ChIP or KAPA HyperPrep were analyzed (using the same parameters) against both reference panels. **A,** The pair of tumor fraction estimations for each sample is shown, where x-axis represents estimation from TruSeq ChIP panel and y-axis represents estimation from KAPA HyperPrep panel. **B,** zoomed-in view on the cfDNA samples with the tumor fraction below 10%. The comparison shows a linear relationship with high concordance between the estimations from different reference panels.

**Supplemental Tables**

**Supplemental Table 1.**

| | Type | Stage | | Total sample | Predicted as malignant | Sensitivity at 98% specificity | 95% CI | ichorCNA TF > 3% |
|---|---|---|---|---|---|---|---|---|
| **Healthy** | | | | 260 | 4 | | | |
| **Tumor** | | | | 238 | 220 | 92.44% | 88.31% - 95.46% | 126 (52.95%) |
| | **HL** | | | 179 | 170 | 94.97% | 90.67% - 97.68% | 93 (51.96%) |
| | | Early stage | | 155 | 148 | 95.48% | 90.92% - 98.17% | 74 (47.74%) |
| | | | I | 10 | 9 | 90.00% | 55.50% - 99.75% | 7 (70.00%) |
| | | | II | 145 | 139 | 95.86% | 91.21% - 98.47% | 67 (46.21%) |
| | | Advanced stage | | 24 | 22 | 91.67% | 73.00% - 98.97% | 19 (79.17%) |
| | | | III | 9 | 9 | 100.00% | 66.37% - 100.00% | 9 (100.00%) |
| | | | IV | 15 | 13 | 86.67% | 59.54% - 98.34% | 10 (66.67%) |
| | **DLBCL** | | | 37 | 32 | 86.49% | 71.23% - 95.46% | 21 (56.76%) |
| | | Early stage | | 6 | 5 | 83.33% | 35.88% - 99.58% | 2 (33.33%) |
| | | | I | 1 | 1 | 100.00% | 2.50% - 100.00% | 1 (100.00%) |
| | | | II | 5 | 4 | 80.00% | 28.36% - 99.49% | 1 (20.00%) |
| | | Advanced stage | | 15 | 13 | 86.67% | 59.54% - 98.34% | 11 (73.33%) |
| | | | III | 7 | 7 | 100.00% | 59.04% - 100.00% | 7 (100.00%) |
| | | | IV | 8 | 6 | 75.00% | 34.91% - 96.81% | 4 (50.00%) |
| | | Unknown | | 16 | 14 | 87.50% | 61.65% - 98.45% | 8 (50.00%) |
| | **MM** | | | 22 | 18 | 81.82% | 59.72% - 94.18% | 12 (54.55%) |
| | | | I | 3 | 3 | 100.00% | 29.24% - 100.00% | 1 (33.33%) |
| | | | II | 7 | 5 | 71.43% | 29.04% - 96.33% | 3 (42.86%) |
| | | | III | 7 | 7 | 100.00% | 59.04% - 100.00% | 7 (100.00%) |
| | | | unknown | 5 | 3 | 60.00% | 14.66% - 94.73% | 1 (20.00%) |

**Supplemental Table 2.**

| | DLBCL | HL | MM |
|---|---|---|---|
| Sensitivity | 0.6563 | 0.9000 | 0.7778 |
| Specificity | 0.9096 | 0.8200 | 0.9703 |
| PPV | 0.5526 | 0.9444 | 0.7000 |
| NPV | 0.9396 | 0.7069 | 0.9800 |
| Prevalence | 0.1455 | 0.7727 | 0.0818 |
| Detection Rate | 0.0955 | 0.6955 | 0.0636 |
| Detection Prevalence | 0.1727 | 0.7364 | 0.0909 |
| Balanced Accuracy | 0.7829 | 0.8600 | 0.8740 |

**Supplemental Table 3.**

| | Type | Stage | | Total Sample | Predicted as malignant | Sensitivity at 95% specificity | 95% CI | ichorCNA TF >3% |
|---|---|---|---|---|---|---|---|---|
| Healthy | | | | 107 | 5 | - | | |
| Malignant | | | | 320 | 177 | 55.31% | 49.68% - 60.84% | 85 (26.56%) |
| | Breast | | | 46 | 7 | 15.22% | 6.34% - 28.87% | 3 (6.52%) |
| | | Early stage | | 35 | 2 | 5.71% | 0.70% - 19.16% | 0 (0.00%) |
| | | | I | 23 | 1 | 4.35% | 0.11% - 21.95% | 0 (0.00%) |
| | | | II | 12 | 1 | 8.33% | 0.21% - 38.48% | 0 (0.00%) |
| | | Advanced stage | | 11 | 5 | 45.45% | 16.75% - 76.62% | 3 (27.27%) |
| | | | III | 5 | 1 | 20.00% | 0.51% - 71.64% | 1 (20.00%) |
| | | | IV | 6 | 4 | 66.67% | 22.28% - 95.67% | 2 (33.33%) |
| | Colorectal | | | 70 | 25 | 35.71% | 24.61% - 48.07% | 10 (14.29%) |
| | | Early stage | | 36 | 13 | 36.11% | 20.82% - 53.78% | 3 (8.33%) |
| | | | I | 19 | 7 | 36.84% | 16.29% - 61.64% | 0 (0%) |
| | | | II | 17 | 6 | 35.29% | 14.21% - 61.67% | 3 (17.65%) |
| | | Advanced stage | | 34 | 12 | 35.29% | 19.75% - 53.51% | 7 (20.59%) |
| | | | III | 25 | 6 | 24.00% | 9.36% - 45.13% | 4 (16.00%) |
| | | | IV | 9 | 6 | 66.67% | 29.93% - 92.51% | 3 (33.33%) |
| | GIST | Advanced stage | | 35 | 28 | 80.00% | 63.06% - 91.56% | 12 (34.29%) |
| | Lung | Advanced stage | | 44 | 30 | 68.18% | 52.42% - 81.39% | 17 (38.64%) |
| | Ovarian | | | 125 | 87 | 69.60% | 60.74% - 77.51% | 43 (34.40%) |
| | | Early Stage | | 36 | 13 | 36.11% | 20.82% - 53.78% | 3 (8.33%) |
| | | | I | 25 | 8 | 32.00% | 14.95% - 53.50% | 2 (8.00%) |
| | | | II | 11 | 5 | 45.45% | 16.75% - 76.62% | 1 (9.09%) |
| | | Advanced stage | | 80 | 68 | 85.00% | 75.26% - 92.00% | 36 (45.00%) |
| | | | III | 49 | 43 | 87.76% | 75.23% to 95.37% | 23 (46.94%) |

| | | | | | | |
|---|---|---|---|---|---|---|
| | IV | 31 | 25 | 80.65% | 62.53% - 92.55% | 13 (41.94%) |
| Metastatic | | 9 | 6 | 66.67% | 29.93% - 92.51% | 4 (44.44%) |

**Supplemental Table 4.**

| | Breast | Colorectal | GIST | Lung | Ovarian |
|---|---|---|---|---|---|
| Sensitivity | 0.0000 | 0.4000 | 0.7143 | 0.6667 | 0.8395 |
| Specificity | 0.9878 | 0.8972 | 0.9790 | 0.9433 | 0.7222 |
| PPV | 0.0000 | 0.4000 | 0.8696 | 0.7143 | 0.7312 |
| NPV | 0.9585 | 0.8972 | 0.9459 | 0.9301 | 0.8333 |
| Prevalence | 0.0409 | 0.1462 | 0.1637 | 0.1754 | 0.4737 |
| Detection Rate | 0.0000 | 0.0584 | 0.1170 | 0.1170 | 0.3977 |
| Detection Prevalence | 0.0117 | 0.1462 | 0.1345 | 0.1637 | 0.5439 |
| Balanced Accuracy | 0.4939 | 0.6486 | 0.8467 | 0.8050 | 0.7809 |
